# Caregiver Needs and Technology Acceptability for Behavioral-Crisis Support in Children With Neurodevelopmental or Behavioral Conditions

**DOI:** 10.64898/2026.07.09.26357685

**Authors:** Fenil Patel, Brittany Williams, Rana Elmaghraby, Ernest V. Pedapati

## Abstract

**Background:** Behavioral crises are common and distressing in children with neurodevelopmental or behavioral conditions, and many escalate to emergency service use. Access to behavioral therapy is often constrained. Smartphone applications, in-home systems, and wearable sensors that could support caregivers during crises at home are in active development, but few studies have asked caregivers what they would accept or want from such tools.

**Methods:** We conducted a single-center cross-sectional online survey (REDCap) of caregivers of children aged 5–17 years with neurodevelopmental or behavioral conditions, recruited as a convenience sample through flyers, email invitations, and in-person invitations during clinic visits from the neurobehavioral continuum of care at Cincinnati Children’s Hospital Medical Center. The response rate is undetermined due to the open-ended recruitment process. Prior behavioral-crisis experience was not an eligibility requirement. The 24-item instrument covered crisis burden, service utilization, caregiver confidence and training, therapy access and barriers, and technology preferences. Analyses were estimation-first (proportions with Wilson 95% confidence intervals [CIs]; medians with interquartile ranges [IQRs]); three pre-specified bivariate analyses used ordinal methods (Kendall’s tau-b and Jonckheere–Terpstra for ordinal pairs; Friedman for repeated ratings of five support functions). Recruitment closed on July 22, 2026 with 84 respondents (one additional study-team test entry was excluded), exceeding the feasibility-based enrollment target of 75; this full-sample analysis supersedes the interim analysis of the first 55 respondents posted as version 1 of this preprint, and all findings remain hypothesis-generating.

**Results:** Seventy-eight of 84 responding caregivers (93%; 95% CI 85–97%) reported that their child had ever experienced a behavioral crisis; 43% (95% CI 33–54%) reported crises at least weekly, and 27% (95% CI 19–38%) had ever used 911 or an emergency department for a crisis. Half of caregivers (52%) felt not at all or only a little confident managing crises, and fewer than half (48%; 95% CI 37–58%) had received informal or formal crisis-management training. The most frequent barrier to behavioral therapy was long waitlists (57%; 95% CI 46–67%). Stated openness to hypothetical technology-based crisis support was high, with 60% (95% CI 49–69%) very interested in a smartphone app or in-home support system, 81% (95% CI 71–88%) willing to have their child use a wearable sensor (1 of 84 declined), and 49% (95% CI 39–60%) willing to share video or audio with a future support tool (a further 43% answered “maybe”; 7% declined). The most-valued features were a personalized crisis plan (63%) and safe de-escalation scripts (46%); the most-cited concern was privacy and data security (38%).

**Conclusions:** In this self-selected, single-center sample, caregivers of children with neurodevelopmental or behavioral conditions reported substantial crisis burden, limited training, and constrained access to therapy, alongside high stated openness to technology-based crisis support; personalization and privacy were their leading priorities. These preliminary, hypothesis-generating findings can inform the design of caregiver-facing crisis-support technologies and larger representative studies.

## Introduction

Behavioral crises, defined here as episodes of severe agitation, aggression, self-injury, or elopement requiring immediate intervention, are common and consequential for children with autism and other neurodevelopmental or behavioral conditions. These behaviors are related but distinct, and each is common. A meta-analysis of 37 studies (N = 14,379) estimated a pooled prevalence of self-injurious behavior of 42% (95% CI 38–47%) among individuals with autism spectrum disorder (ASD)^1^, and aggression toward caregivers has been reported in 68% of children with ASD^2^. Elopement affects roughly one in four children with developmental disabilities in nationally representative data^3^, often recurrently and with substantial family disruption^4^. In a survey of 462 parents of youth with ASD, 32% reported a mental-health crisis in the preceding three months, with associated parental depression and reduced family quality of life^5^. Most of this evidence comes from children with autism or intellectual and developmental disabilities (IDD), the populations in which crises are best characterized. The present survey, like the clinical settings it drew from, spans a broader neurodevelopmental and behavioral population.

Families facing these crises frequently turn to emergency systems. Children with ASD are approximately nine times more likely than peers to have a psychiatric-related emergency department (ED) visit^6^, and children with IDD use ED and inpatient care at 1.8 times the rate of the general pediatric population, roughly doubling annual costs^7^. Emergency contact, moreover, does not resolve these crises. In a 10-year review of one psychiatric emergency department, patients with ASD or intellectual disability presented with high acuity and were often difficult to transition to inpatient or other definitive treatment settings^8^, and in a survey of 2,525 caregivers, crisis events were often associated with law-enforcement contact and psychiatric hospitalization^9^.

Behavioral therapy and structured parent training are the mainstay upstream response, with the strongest evidence for disruptive behavior in ASD and related conditions^10–12^. Both are effective but structurally difficult to access. More than half of United States counties have no Board Certified Behavior Analyst, and workforce growth has not eliminated geographic inequity^13–15^. Coverage type further shapes access and out-of-pocket burden^16^, and children with ASD are more likely than other children with special healthcare needs to report unmet therapy need (odds ratio [OR] 1.42; 95% CI 1.18–1.71)^17^. Caregivers, meanwhile, do most of the day-to-day management. Low parental self-efficacy is consistently linked to stress and poorer mental health^18,19^. Parent training raises self-efficacy (standardized mean difference 0.60 across 25 randomized trials^20^) and reduces disruptive behavior in randomized trials^10–12^, but benefits are not universal and are hard to deliver at scale. Caregiver strain persists over years^21^, and one large trial of adapted parent training (Stepping Stones Triple P) in preschoolers with moderate-to-severe intellectual disability found no significant benefit on its primary outcome, parent-reported child behavior^22^.

Technology may help address part of this problem at home. Parent-facing mobile health (mHealth) applications and web-based training are feasible and show preliminary benefit for parenting stress and child behavior in small, largely pilot-stage studies^23–27^. Real-time support is beginning to reach trials. Just-in-time adaptive interventions provide a framework for delivering support at the moment a person’s state calls for it^28,29^. A smartwatch-augmented parent–child interaction therapy trial delivered heart-rate-triggered prompts with high adherence^30^, and artificial intelligence (AI)-based systems are under active development, from escalation-decision support to predictive caregiver-alerting and applied-behavior-analysis platforms and, most recently, large-language-model parent-support agents^31–34^. Wearable physiological sensing has anticipated behavioral escalation about a minute before it becomes observable, although largely in inpatient samples of youth with ASD rather than in homes^35–38^. There are methodological limitations to these studies, as flagged by a recent systematic review^39^.

This literature, however, is almost entirely technology-feasibility, prediction, and clinician-or researcher-facing. To our knowledge, based on structured though not systematic searches of PubMed and the reference lists of recent reviews, no published study has quantified what caregivers of children with neurodevelopmental or behavioral conditions find acceptable, want, and worry about in technology-based crisis support at home. As a result, these systems are being designed and moved toward trials without a quantitative read on whether the caregivers who would use them at home would accept them, or on which features matter most to those caregivers. The present survey addresses this question directly, from the demand side.

We therefore conducted a cross-sectional survey of caregivers of children aged 5–17 years with neurodevelopmental or behavioral conditions with four objectives: (1) to assess caregiver confidence and training in managing behavioral crises; (2) to identify challenges and service utilization during behavioral crises; (3) to identify barriers to accessing behavioral therapy services; and (4) to evaluate acceptability of technology-based crisis-support systems and identify priority features. Although recruitment closed above the protocol target, the sample remains modest, single-center, and self-selected, so we report these results as an explicitly hypothesis-generating needs assessment.

## Methods

### Study design and setting

We conducted a cross-sectional online survey using REDCap electronic data capture tools^40^ hosted on Cincinnati Children’s Hospital Medical Center (CCHMC) secure servers. The study was approved by the CCHMC Institutional Review Board (#2025-0646; expedited category 7) and is reported in accordance with the STROBE statement for cross-sectional studies^41^ and the CHERRIES checklist for internet e-surveys^42,43^ (completed checklists in the Supplement). The survey opened on December 29, 2025 and closed on July 22, 2026; it received 85 submissions in that window, of which 84 were caregiver responses (one study-team test entry was excluded; see Data handling). Version 1 of this preprint reported an interim analysis of the first 55 responses (received through June 26, 2026); this full-sample version supersedes it.

### Participants and recruitment

Eligible respondents were parents or legal caregivers, aged 18 years or older and able to complete a survey in English, of a child aged 5–17 years with a neurodevelopmental and/or behavioral condition. Caregivers were recruited from the neurobehavioral continuum of care at CCHMC through three channels: recruitment flyers, email invitations, and in-person invitations during clinic visits; the survey was not advertised on the open web. Recruitment materials presented the study as a survey about supporting families during behavioral crises. Recruitment materials mentioned a raffle for one of ten $25 gift cards, for which only completed surveys were eligible. Participation was voluntary. Access to the questionnaire was gated by a three-item eligibility screen (caregiver age, child age/condition, and English ability). Eligible caregivers then viewed a study-information screen presenting the study purpose and procedures, voluntary participation, risks and benefits, confidentiality protections, and contact information, and had to click “I AGREE” before accessing the questions, constituting implied consent for this minimal-risk study. Contact information provided for the raffle was stored separately from survey responses. This was a convenience sample of caregivers already engaged with CCHMC’s neurobehavioral services; prior behavioral-crisis experience was not an eligibility criterion. Because recruitment combined flyers, email, and in-person invitation, the number of caregivers reached cannot be ascertained; view and participation rates therefore cannot be computed, and counts of caregivers who opened the survey or failed the eligibility screen were not retained by the survey platform. Safeguards against duplicate entry were limited to the clinic-anchored recruitment frame and the separation of raffle contact information from responses; no IP-based or cookie-based duplicate screening was performed.

### Survey instrument

The instrument comprised 24 items in five sections: About Your Child (4 items), Your Child’s Behaviors and Characteristics (5 items), Your Experience and Support (4 items + 1 conditional sub-item), Treatment History and Access (4 items + 1 conditional sub-item), and Technology Solution Preferences (7 items). Items were developed de novo by the study team for this study and were not adapted from existing validated measures; key constructs (crisis frequency, confidence, training, interest, willingness, concerns) were each measured with a single item. The questionnaire embedded a caregiver-facing, study-specific definition of a behavioral crisis, displayed alongside the crisis items: “an episode where your child’s behavior becomes intense, unsafe, or disruptive enough that you need to act right away,” with concrete examples (hitting, self-injury, breaking things, running off, or a meltdown that will not stop). Item formats were single-response (radio/yes-no), multi-response (checkbox; two items capped at three selections), and limited free text. Estimated completion time was 10–15 minutes. Before launch, the instrument was pre-tested by clinical colleagues for functionality and user experience; it was not piloted with caregiver families. The survey was administered over eight screens: an eligibility screen, a study-information/consent screen, one screen per instrument section (five), and a final thank-you screen with a link to the separate raffle entry. The three eligibility items, the consent response, and the child’s age were required fields; all other items were optional. Respondents could return to previous screens to change answers and could save and return later to complete the survey, but answers could not be altered after final submission. Items were not randomized; REDCap displayed conditional sub-items (training helpfulness; therapy types) only to respondents meeting the skip-logic criteria. The full instrument is provided as a supplementary appendix.

### Variables and denominators

Analysis variables followed the REDCap data dictionary. Two conditional items used restricted denominators enforced throughout: training helpfulness was asked only of respondents reporting informal or formal training, and therapy types only of respondents whose child ever received behavioral therapy. For each item, respondents who skipped it were excluded from that item’s denominator, and the N used is reported alongside every estimate. Derived variables (e.g., any informal/formal training; weekly-or-more crisis frequency; any 911/ED use; any outside help during a crisis) were defined in a documented codebook before the descriptive analysis. The crisis-actions item asked about actions ever taken “in the past, during a crisis”; it therefore reflects lifetime utilization, not per-episode rates. ZIP code was collected but is not reported here to protect against re-identification of a small sample; parent-reported diagnoses are summarized descriptively and were not used as clinical covariates.

### Data handling and missing data

Analyses used a de-identified export. Free-text fields were reviewed only in aggregate and are not quoted. The export contained 85 records; record review identified one as a test entry made by the study team (it contained only the eligibility and consent screens and no survey responses), and it was excluded, leaving 84 respondents. Of these, 83 completed the survey and 1 partially completed it; the partial record was included in descriptive analyses for the items it answered and its exclusion was checked as a sensitivity analysis (no headline proportion changed by more than 0.9 percentage points, and no interpretation changed). Item-level missingness on always-shown items was low (at most 2 of 84 on any single item); apparent missingness above 50% occurred only for skip-logic sub-items and optional free-text fields, as expected by design. Analyses were complete-case per item or per analysis, with the N used reported for each.

### Statistical methods

The analysis was estimation-first: proportions are reported with Wilson 95% CIs, distributions as medians with IQRs, and association strengths as effect sizes with 95% CIs; p-values are secondary and no result is treated as confirmatory. For multi-response items, percentages use respondents (not responses) as the denominator and therefore do not sum to 100%.

Three bivariate analyses were pre-specified in a written analysis plan (see the Transparency statement below for its timing and revisions): (A1) crisis-management training (ordinal, 4 levels) versus confidence managing crises (ordinal, 4 levels), analyzed with Kendall’s tau-b (95% CI) and the Jonckheere–Terpstra trend test, with a collapsed two-level (any informal/formal training vs. none/some guidance) Wilcoxon rank-sum comparison and a Monte-Carlo Fisher’s exact test (B = 10,000) as sensitivity analyses; (A2) rated helpfulness of five hypothetical support functions (5-point ordinal, repeated measures), analyzed with the Friedman omnibus test and Kendall’s W among complete cases, with Holm-adjusted pairwise Wilcoxon signed-rank tests planned only if the omnibus was significant; and (A3) caregiver education (ordinal, 4 levels) versus interest in a smartphone app (ordinal, 3 levels), analyzed as in A1. Additional exploratory associations are labeled as such and were not corrected for multiplicity; they should be interpreted only as context for future hypotheses.

Analyses were performed in R 4.5.2 (tidyverse, gtsummary, janitor, naniar, DescTools, clinfun, binom) with a fixed random seed; key statistics were independently recomputed in Python 3.14 (pandas, SciPy, statsmodels) and matched. All tables and figures are generated by code from the analysis outputs, and the full pipeline passed a clean-room reproducibility test (all output tables byte-identical on re-render from the raw export).

### Transparency statement

The analysis plan was drafted a priori but during data collection (after approximately 20 of the eventual 84 caregiver responses had been received) and was not publicly pre-registered. That plan fixed the three comparisons (A1–A3). Before any inferential analysis was conducted, the originally planned chi-square tests were replaced, in a dated written decision made on distributional grounds (sparse tables; ordinal variables), by the ordinal methods reported here, with the collapsed two-level comparison and Fisher’s exact tests retained as sensitivity analyses; the exploratory analyses (E1–E3) were specified in the same written plan, also before the inferential analyses were run. An interim analysis of the first 55 responses was posted as version 1 of this preprint; the decision to report it preceded the analyses. Recruitment closed on July 22, 2026 with 84 caregiver responses, exceeding the protocol target of 75, a feasibility-based enrollment target reflecting the anticipated recruitment window and clinic volume rather than a formal power or precision calculation; for context, a proportion of 50% carries a Wilson 95% CI half-width of approximately ±11 percentage points at N = 84. The study remains underpowered for its pre-specified comparisons, and we frame every result as preliminary and hypothesis-generating.

## Results

### Participants

The survey received 85 submissions; after exclusion of the study-team test entry, 84 caregivers were included, of whom 83 (99%) completed the survey and 1 partially completed it (included where data were present). Because recruitment combined flyers, email invitations, and in-person clinic invitations, the number of caregivers reached is unknown and a response or view rate cannot be computed. Sample characteristics are shown in **Table 1**. The median child age was 10 years (IQR 8–13.25; range 5–17). Caregivers most often identified their child as White (82%); 21% identified their child as Black/African American. Children carried a median of 3 named diagnoses (IQR 2–4). The most common were attention-deficit/hyperactivity disorder (81%; 95% CI 71–88%), anxiety disorder (51%; 95% CI 41–62%), and ASD (50%; 95% CI 40–60%); 35% had an intellectual disability or developmental delay. Ten percent of children had limited or no functional communication, and 31% were taking four or more behavioral/mental-health medications. Nearly all households (99%) had WiFi or broadband internet; 3 of 84 caregivers (3.6%) reported limited or unreliable service, and none reported having no home internet access.

**Table 1:**
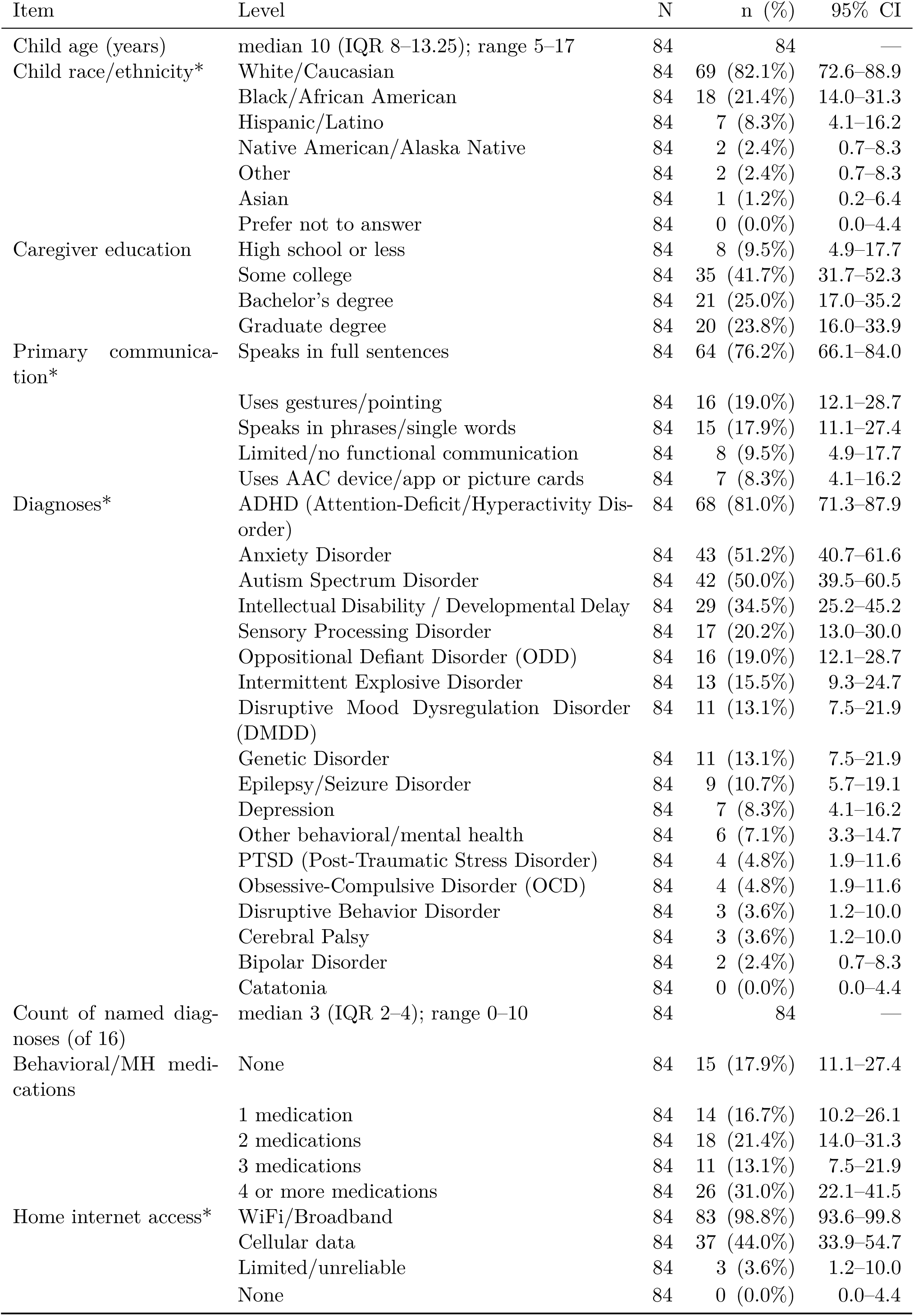
Sample characteristics (N = 84; the N column gives each item’s denominator). Multi-response items (*) allow multiple selections per respondent, so percentages do not sum to 100%. CI = Wilson 95% confidence interval.

### Crisis burden and service utilization

Nearly every respondent (78/84; 93%; 95% CI 85–97%) reported that their child had ever experienced a behavioral crisis, although crisis history was not an eligibility requirement; 5 answered no and 1 was unsure. This near-universal history is consistent with sampling from clinics where crises are common and with self-selection of crisis-affected families into the survey. One respondent reported that crises no longer occurred. Crises occurred at least weekly for 43% of families (95% CI 33–54%), including daily crises for 12% (**Table 2**). The most frequently selected challenging behaviors (up to three selections) were meltdowns (62%), hitting/kicking/biting (48%), and refusal to follow directions (38%); 27% selected self-injurious behavior.

**Table 2:**
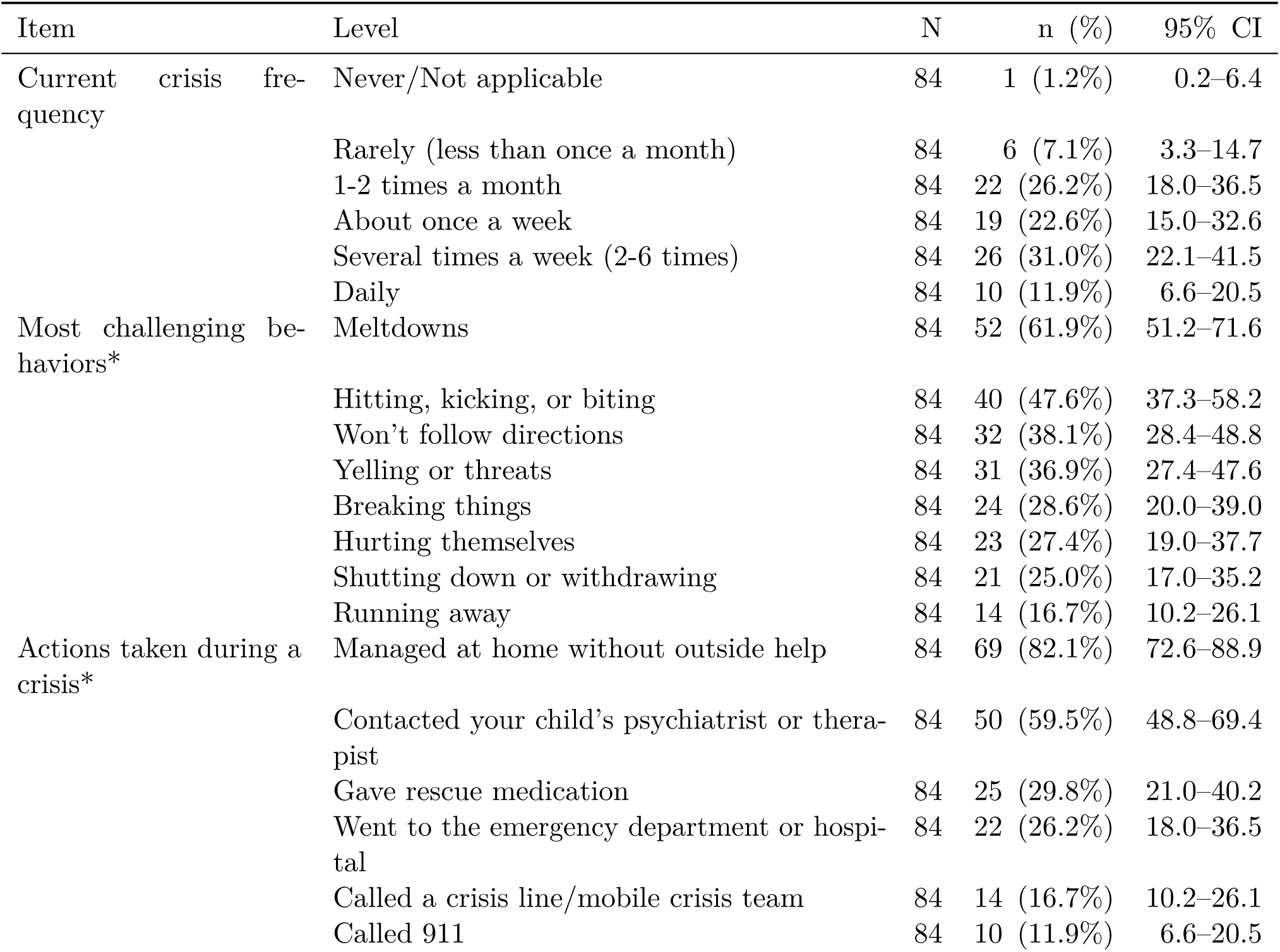
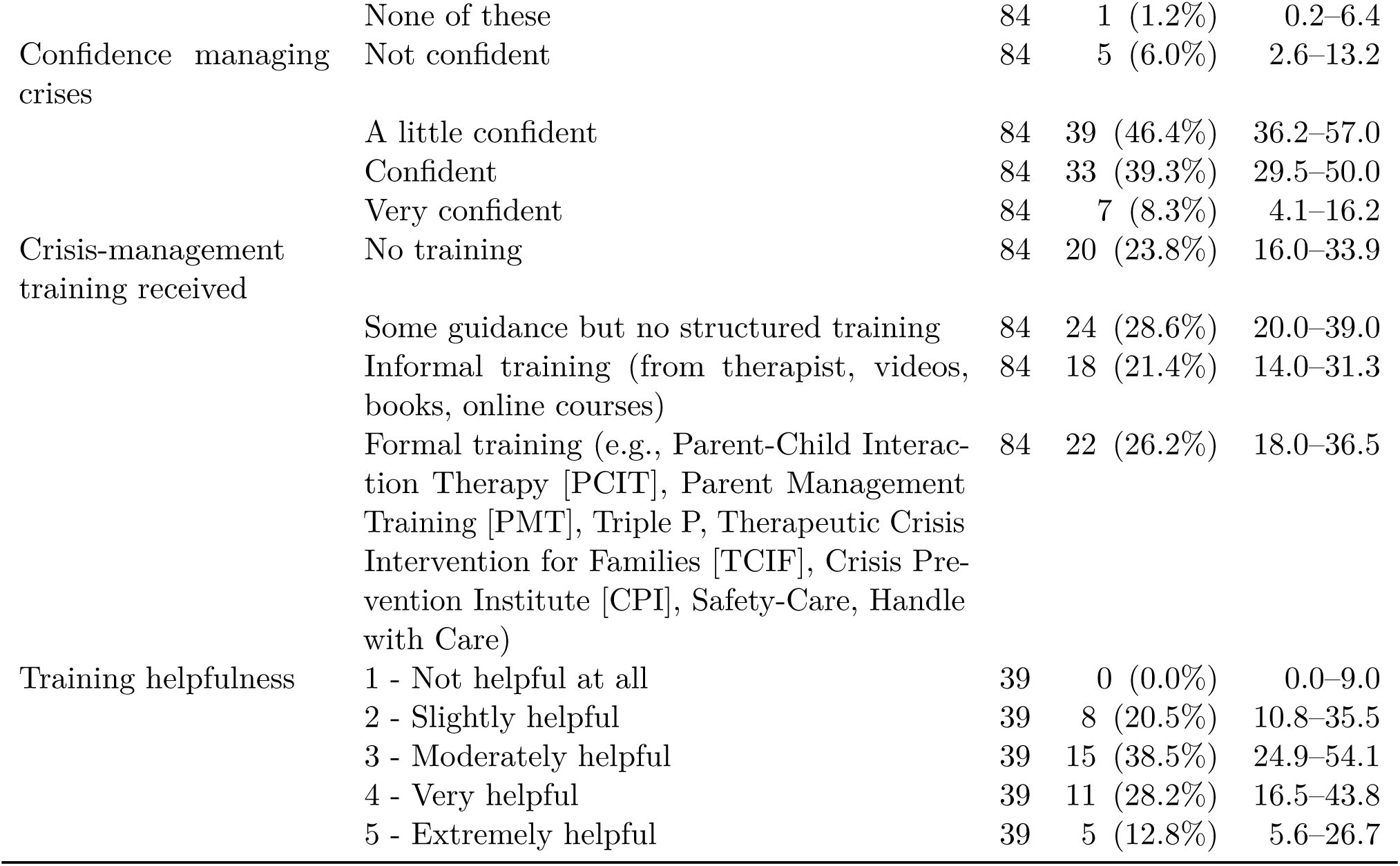
Crisis burden, service utilization, and caregiver confidence and training (N = 84; the N column gives each item’s denominator). Multi-response items (*) allow multiple selections. The crisis-actions item asked about actions ever taken during a crisis (lifetime, not per-episode). Training helpfulness was asked only of caregivers reporting informal or formal training (n = 39 of 40 eligible answered). CI = Wilson 95% confidence interval.

Most caregivers (82%) had managed at least one crisis at home without outside help, and about two-thirds (65%; 95% CI 55–75%) had ever sought outside help during a crisis. Among all respondents, 60% had contacted the child’s psychiatrist or therapist, 30% had given rescue medication, and 27% (95% CI 19–38%) had used 911 or an ED (26% ED; 12% 911). One respondent selected “none of these.”

### Caregiver confidence and training

Half of caregivers (52%; 44/84) reported feeling not at all confident (6%) or only a little confident (46%) managing their child’s crises; 8% felt very confident. Fewer than half (48%; 95% CI 37–58%) had received informal (21%) or formal (26%) crisis-management training; 24% had no training of any kind and 29% had received only unstructured guidance. Among the 39 trained caregivers who rated their training, 41% (16/39) found it very or extremely helpful and 38% moderately helpful, with 21% rating it only slightly helpful.

### Therapy access and barriers

Most children (82%; 95% CI 73–89%) had ever received behavioral therapy (51% currently; 31% in the past); 10% had never sought services and 3 children (3.6%) were on a waitlist at the time of the survey. Among ever-treated children (n = 69), the most common services were general behavioral therapy (54%), talk therapy such as cognitive-behavioral therapy (46%), and autism-focused therapy such as applied behavior analysis (32%).

Most caregivers reported barriers. The median number reported was 2 (IQR 1.75–4; range 0–9), and only 12% (95% CI 7–21%) reported no barriers and good access (**Figure 1**). The leading barriers were long waitlists (57%; 95% CI 46–67%), therapy times conflicting with work or school (44%; 95% CI 34–55%), and insurance not covering or limiting coverage (42%; 95% CI 32–52%). (The barrier item asked about barriers ever faced in accessing therapy; therapy status reflects the time of the survey.) Notably, 18% reported no qualified providers in their area, whereas transportation (5%) and language (1%) barriers were rare in this sample.

**Figure 1:**
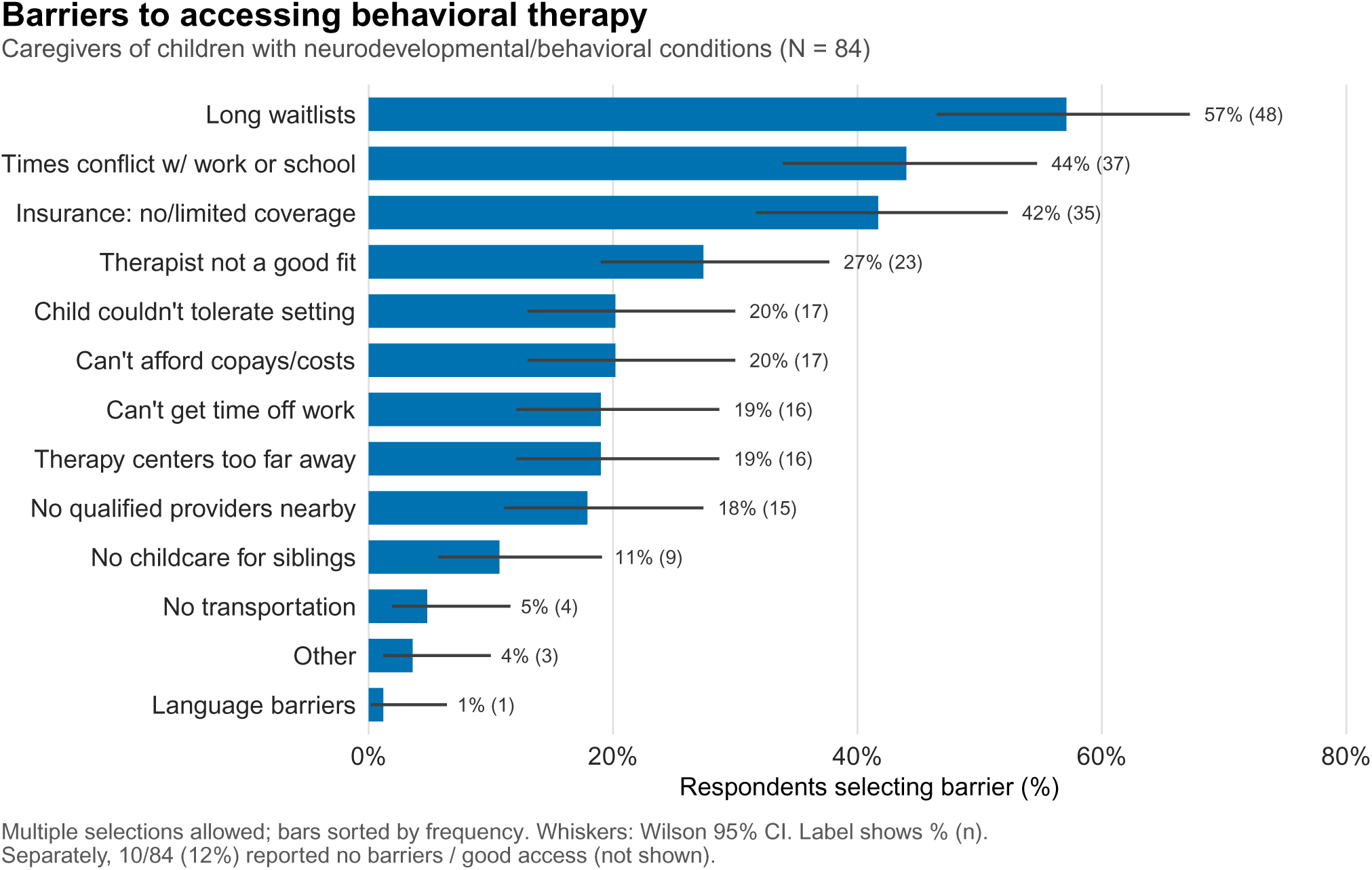
Caregiver-reported barriers to accessing behavioral therapy (N = 84). Each bar shows the percentage of all respondents who selected a given barrier. Respondents could select multiple barriers, so percentages do not sum to 100%. Bars are sorted by frequency; whiskers are Wilson 95% CIs; data labels show the percentage and (count). Separately, 10 of 84 caregivers (12%) reported no barriers and good access (not shown).

### Technology preferences and acceptability

Stated openness to technology-based crisis support was high (**Figure 3**; exact counts and CIs for all technology items are provided in **Supplementary Table S1**). A majority of caregivers (43 of 84 [51%]; 95% CI 41–62%) were very interested in a smartphone app for crisis support, and a further 43% were somewhat interested; only 6% were not interested. Interest in an in-home camera/speaker system was more divided (23 of 82 [28%] very interested; 25 of 82 [30%] not interested). Overall, 50 of 84 (60%; 95% CI 49–69%) were very interested in at least one of the two modalities.

Sixty-eight of 84 caregivers (81%; 95% CI 71–88%) would let their child wear a physiological sensor such as a heart-rate or skin-temperature monitor; 15 answered “maybe” and only 1 (1.2%) declined outright. Forty-one of 83 (49%; 95% CI 39–60%) would share video or audio with a future support tool, with a further 36 (43%) answering “maybe” (6 [7%] declined). Nearly all caregivers were at least somewhat familiar with AI (83 of 84 [99%]; 61% very familiar).

The most-valued features (up to three selections; **Figure 2**) were a crisis plan personalized to the child (63%; 95% CI 52–73%), safe de-escalation scripts and visuals (46%; 95% CI 36–57%), and step-by-step guidance during a crisis (45%; 95% CI 35–56%), followed by tools to track triggers and early-warning signs (40%). Live support from a trained helper (35%), sharing updates with the care team (25%), and post-crisis debriefing (17%) ranked lower. The most frequently cited greatest concern was privacy or data security (38%; 95% CI 28–49%), followed by lack of human connection (18%), worry that the technology would not understand the child’s unique needs (17%), and inaccurate or unhelpful recommendations (15%); 8% had no concerns.

**Figure 2:**
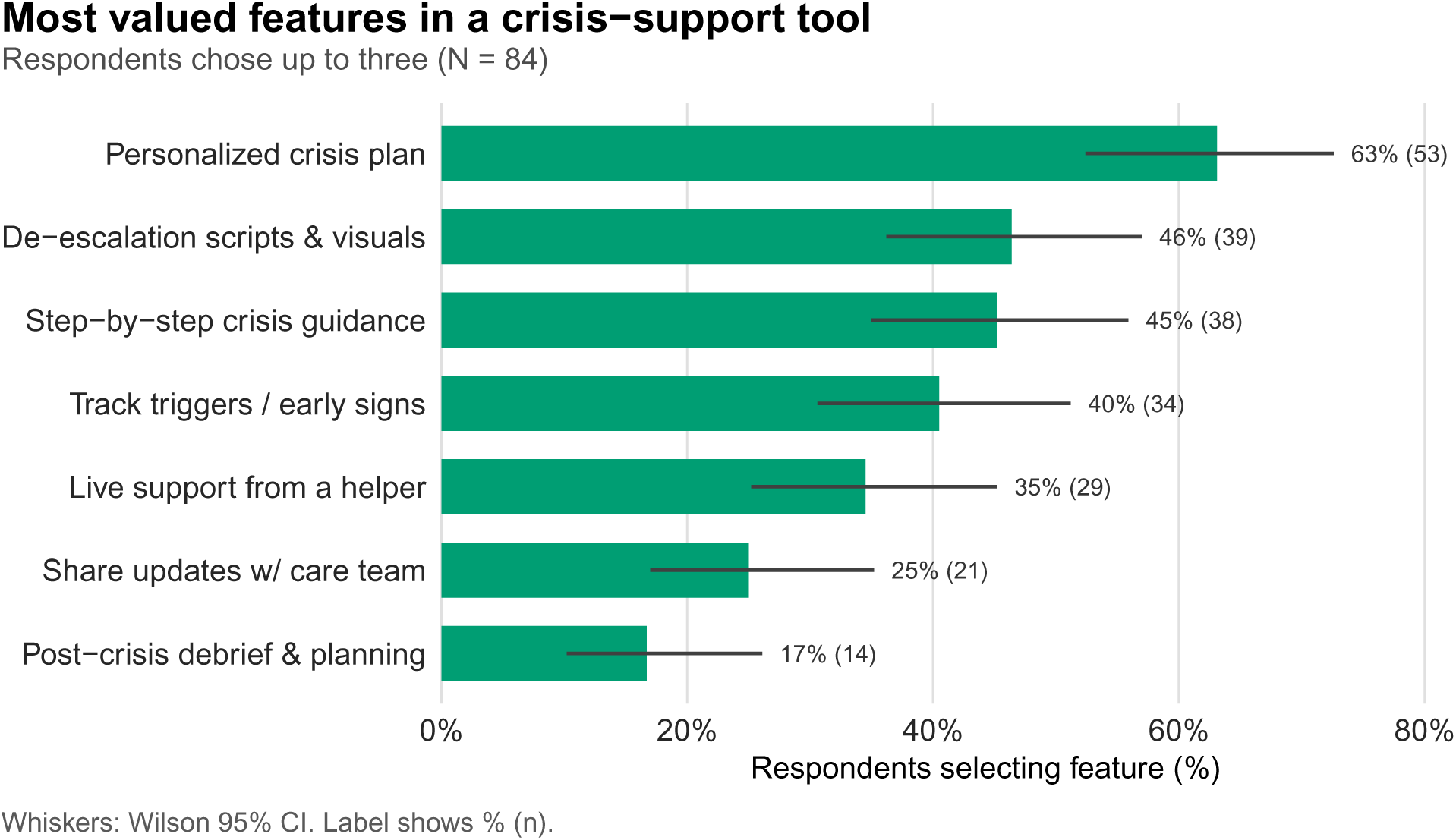
Caregiver-prioritized features for a technology-based crisis-support tool (N = 84). Respondents selected up to three features they would find most valuable; bars show the percentage of all respondents selecting each feature, so percentages do not sum to 100%. Whiskers are Wilson 95% CIs; data labels show the percentage and (count).

**Figure 3:**
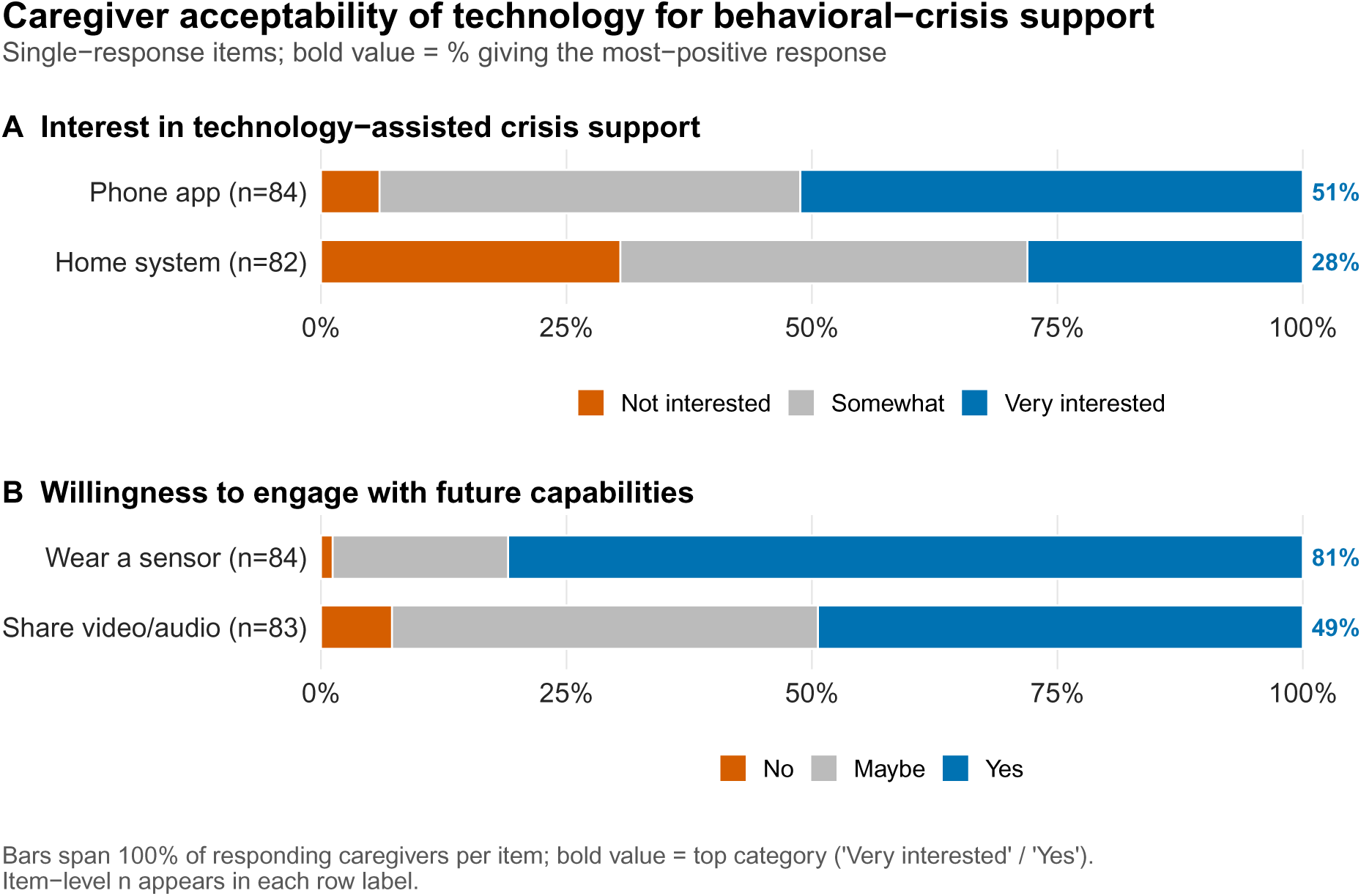
Caregiver acceptability of technology-based behavioral-crisis support. **(A)** Interest in a smartphone app (n = 84) and an in-home camera/speaker system (n = 82). **(B)** Willingness to share video or audio with a future support tool (n = 83) and to have the child wear a physiological sensor (n = 84). Each bar spans 100% of responding caregivers for that item; segments are ordered from the least-to the most-positive response, and the bold value at the right gives the percentage choosing the most-positive response (“Very interested” or “Yes”).

### Pre-specified bivariate analyses (hypothesis-generating)

Results of the three pre-specified analyses are summarized in **Table 3**. Training level and confidence were weakly positively associated (A1: Kendall’s tau-b = 0.16; 95% CI −0.03 to 0.35; Jonckheere–Terpstra p = 0.13), an estimate whose interval includes zero. In the collapsed sensitivity comparison, caregivers with any informal or formal training reported higher confidence than those with none or unstructured guidance only (Wilcoxon rank-sum p = 0.035); the Fisher’s exact sensitivity test on the full table was non-significant (p = 0.28). Given the imprecise primary estimate and the inconsistent sensitivity results, we do not interpret the collapsed comparison as evidence of a training effect.

**Table 3:**
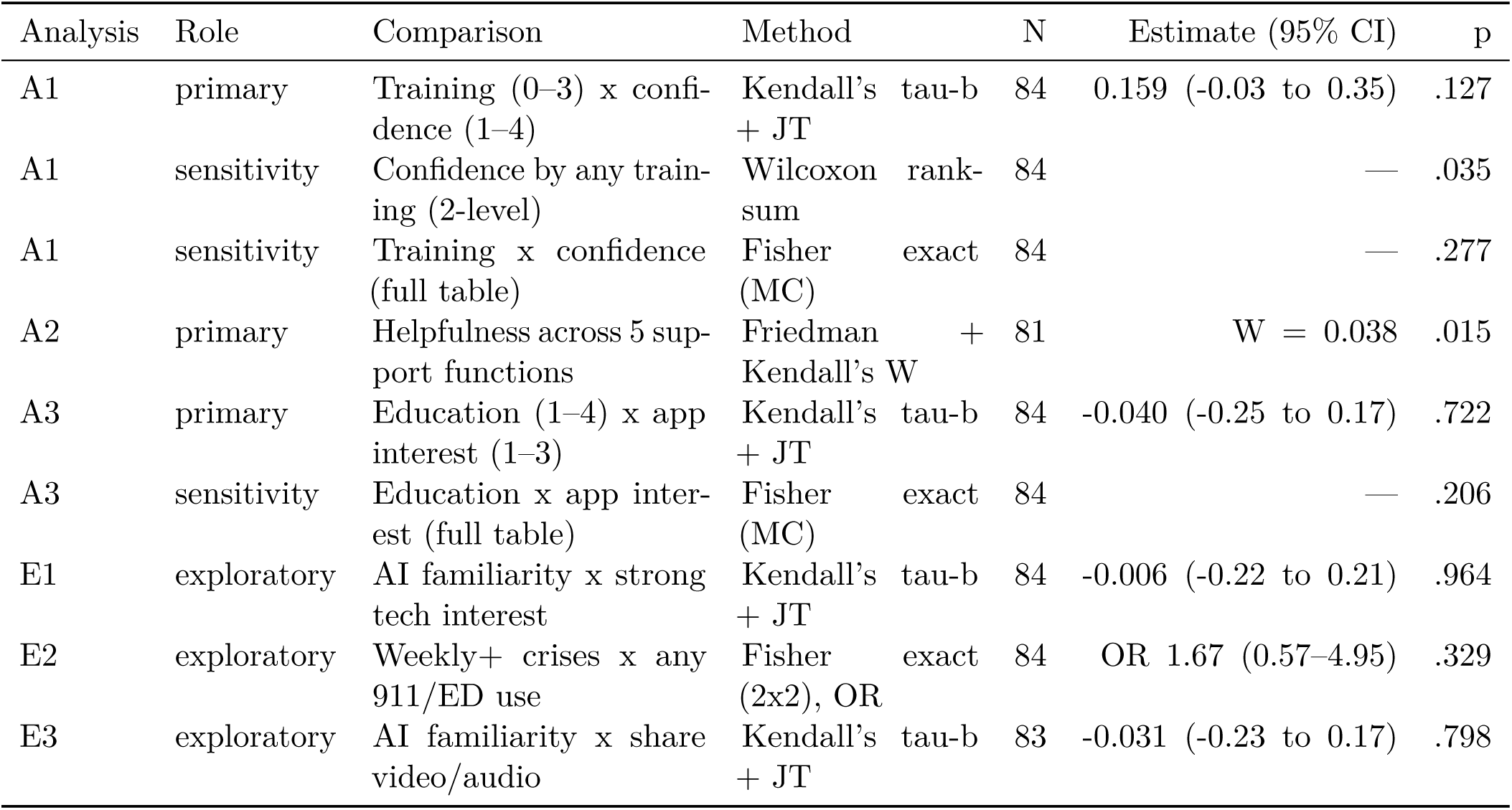
Pre-specified bivariate analyses (A1–A3, with their sensitivity analyses) and clearly-labeled exploratory analyses (E1–E3: not pre-specified, not multiplicity-corrected). Estimates are Kendall’s tau-b with 95% CI unless noted (A2: Kendall’s W; E2: odds ratio). JT = Jonckheere–Terpstra; MC = Monte-Carlo (B = 10,000).

Across the five hypothetical support functions (A2; n = 81 complete cases; 3 of 84 caregivers lacked at least one rating), ratings showed a ceiling effect, with every function receiving a median rating of 4 (“very helpful”) of 5. The Friedman omnibus test was significant (*χ*^2^(4) = 12.37, p = 0.015), but the concordance effect size was trivial (Kendall’s W = 0.04), and in the planned Holm-adjusted post-hoc pairwise comparisons no pair remained significant (smallest adjusted p = 0.11, for the personalized plan versus someone available in the moment; matched-pair r = 0.28). Descriptively, a personalized plan was rated most favorably (mean rank 3.29; 68% very/extremely helpful) and someone available in the moment least (mean rank 2.65; 59% very/extremely helpful) (**Figure 4**).

**Figure 4:**
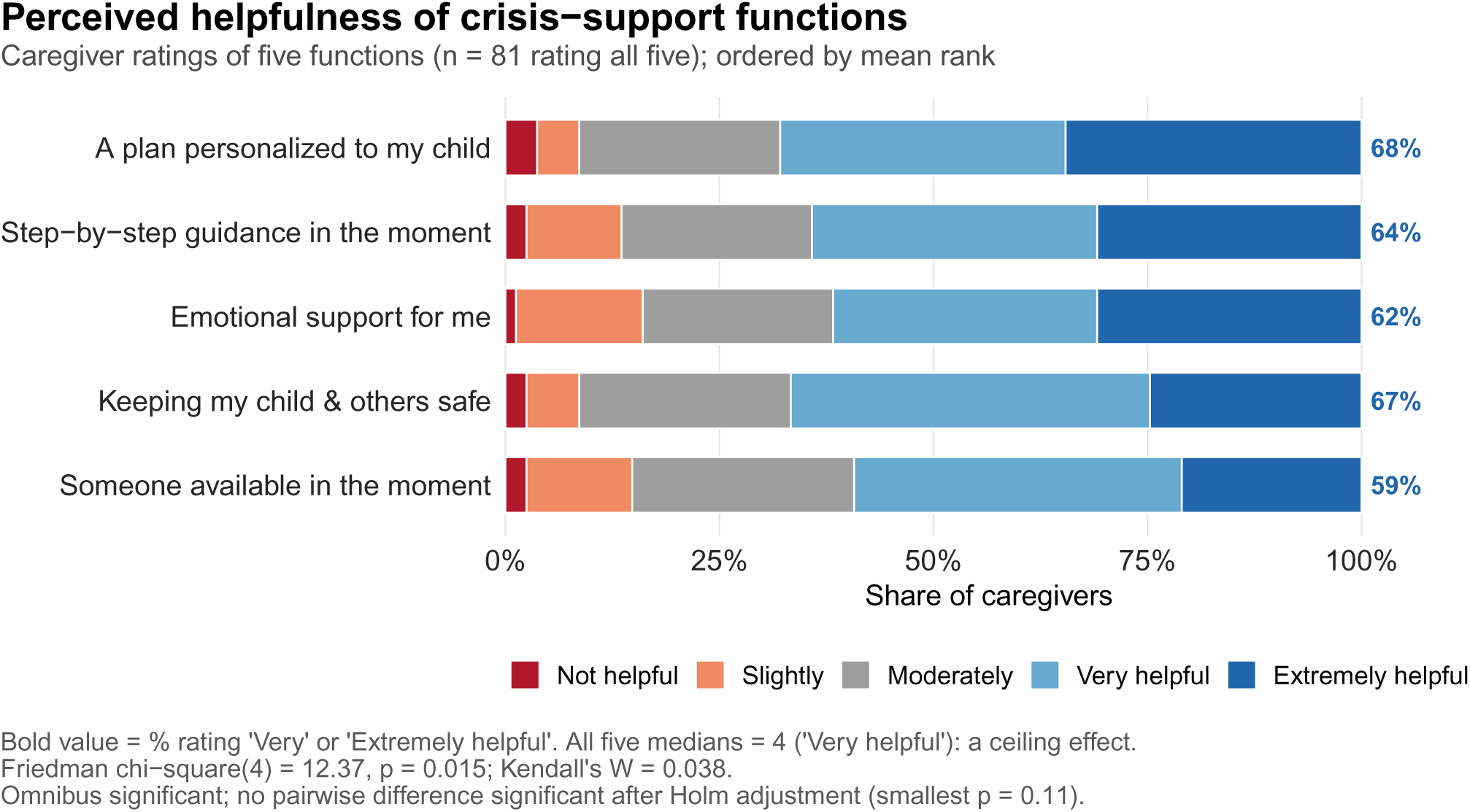
Caregiver-rated helpfulness of five crisis-support functions (n = 81). Distribution of helpfulness ratings on a 5-point scale for five hypothetical support functions, restricted to the 81 caregivers who rated all five (complete-case). Functions are ordered top-to-bottom by mean rank; the bold value at the right is the percentage rating the function “Very” or “Extremely helpful.” All five medians were 4 of 5 (ceiling effect). The Friedman omnibus test was significant (*χ*^2^(4) = 12.37, p =.015) with a trivial concordance effect size (Kendall’s W = 0.04); in the pre-specified Holm-adjusted post-hoc comparisons, no pairwise difference remained significant (smallest adjusted p =.11).

Caregiver education showed no detectable association with interest in a smartphone app (A3: tau-b = −0.04; 95% CI −0.25 to 0.17; p = 0.72); the interval is wide, and these data neither demonstrate nor rule out education-related differences in interest.

### Exploratory analyses

Exploratory associations (Table 3, rows E1–E3; N = 83–84) were imprecise, as expected at this sample size, with confidence intervals compatible with both no association and clinically relevant associations. AI familiarity showed no clear association with strong technology interest (tau-b = −0.01; 95% CI −0.22 to 0.21) or with willingness to share video/audio (tau-b = −0.03; 95% CI −0.23 to 0.17), and weekly-or-more crisis frequency was not significantly associated with 911/ED use (OR 1.67; 95% CI 0.57–4.95).

## Discussion

### Principal findings

In this single-center survey of 84 caregivers of children with neurodevelopmental or behavioral conditions, nearly all of whom reported crisis experience, three findings stand out. First, the crisis burden was heavy and largely home-managed. Crises occurred at least weekly in 43% of families, yet half of caregivers felt little or no confidence managing them and fewer than half had received any structured or informal training. Second, access to the standard of care was constrained. More than half of families faced long waitlists, and 42% reported that insurance did not cover or limited coverage of therapy. This is consistent, at the family level, with structural access problems documented nationally. Third, caregivers in this sample reported high stated openness to hypothetical technology-based crisis support. Three in five were very interested in an app or in-home system, 81% would try a wearable sensor, and only 7% ruled out sharing video or audio (49% yes; 43% “maybe”). Their priorities were specific, led by a plan personalized to their child and concrete de-escalation help, and their leading concern was privacy.

### Comparison with prior work

Our burden and utilization estimates are consistent with the literature that frames behavioral crises as common, consequential events^1,2,5^ that escalate to emergency and law-enforcement systems^6–9^. In our sample, about one in four families had used 911 or an ED for a behavioral crisis. Our barrier profile (waitlists, scheduling conflicts, insurance limits, and provider scarcity) mirrors national workforce and coverage analyses^13–17^. The confidence–training gap we observed is likewise concordant with evidence that parent training improves caregiver self-efficacy^20^ and child behavior^10–12^ but is neither universally effective^22^ nor sufficient to relieve sustained caregiver strain^18,21^. In our data, trained caregivers tended to report higher confidence, which is directionally consistent with that literature, but our study cannot show that training produced the difference. The primary estimate was weak and imprecise (Kendall’s tau-b = 0.16; 95% CI −0.03 to 0.35), meaning the data are compatible with anything from no association to a moderate one. A simpler trained-versus-untrained comparison did reach nominal significance, but it was a sensitivity analysis, not the primary test. And because the survey is cross-sectional, more proactive families may simply be likelier both to seek training and to feel confident, or their crises may differ in severity and resources.

What this study adds is a first quantitative read on caregiver demand. Feasibility-oriented work has established that apps can support parents^23,24^, that real-time physiological sensing can anticipate escalation^35–37,39^, and that just-in-time and AI-augmented supports can be delivered^28,30,31,34^. What that literature has largely not quantified is whether caregivers want these systems in their homes. Our results provide a preliminary, sample-specific answer. Openness to technology-based crisis support was high, and willingness to at least consider a wearable (83 of 84 caregivers; 99%; 95% CI 94–100%) is notable because wearables are the sensing layer of most crisis-prediction prototypes. Willingness expressed for a hypothetical device may still exceed real-world adoption. This aligns with the one qualitative study we identified, in which parents of children with neurodevelopmental disorders were willing to use wearables if feasible and asked for shared design decisions^44^. In our data, caregiver priorities were also specific and design-relevant. Personalization ranked first both prospectively (63% chose a personalized crisis plan as a top-three feature) and in the helpfulness ratings (highest mean rank; 68% very/extremely helpful). This is a hypothesis-generating signal, not a tested comparison, but it suggests generic content may not be enough.

Privacy concerns were equally prominent. Privacy or data security was the most-cited greatest concern (38%), and video/audio sharing drew far more hesitation (43% “maybe”) than wearable sensing. This is consistent with a body of work showing parents are cautious but conditionally willing to accept monitoring technologies, participating mainly when data are private, anonymized, or under their control^45–48^, and with documented confidentiality gaps in child mental-health apps^49^. One interpretation, which future co-design and preference-elicitation studies could test, is a staged rollout that begins with caregiver-controlled, on-device, or wearable-based sensing and personalized guidance and offers continuous audio or video capture only as an opt-in addition once a tool has earned trust.

Finally, remote delivery is plausible in principle. Telehealth-delivered behavioral treatments (parent-implemented intervention and functional-analysis coaching in specific diagnostic groups) have achieved large reductions in problem behavior in trials^50,51^, including in rural settings^52^, although technology-mediated crisis detection and real-time coaching remain untested. The digital divide, however, constrains who can benefit. Rural, uninsured, and Medicaid-insured populations use telehealth the least^53,54^. Minoritized children with ASD have less geographic access to autism resources in the first place^55,56^, on top of pervasive disparities in service use^57,58^. And rural, low-income counties remain short of child-psychiatric care even after accounting for telehealth^59^. Policy levers may help. In observational data, Medicaid home-and community-based waivers were associated with roughly half the unmet need among Black children with ASD^60^. Our sample’s near-universal connectivity (99% WiFi/broadband; no household without internet access; 3.6% with unreliable service) should therefore be read as a feature of a self-selected online sample, not as evidence that connectivity is a solved problem in the catchment population.

### Limitations

This study has important limitations. First, the sample is modest (N = 84, above the feasibility-based protocol target of 75), single-center, and self-selected through an online survey of families already connected to a neurobehavioral continuum of care. The near-universal crisis history (93%) likely reflects three converging influences: the crisis-enriched clinical population sampled, recruitment materials that themselves foregrounded behavioral-crisis support, and selective response by crisis-affected families. As a result, these estimates, particularly the high reported technology openness, may not generalize to less-connected, less crisis-burdened, or less digitally engaged families. Second, all measures are caregiver self-report, including diagnoses, and are subject to recall and social-desirability bias. The crisis-actions item measured lifetime, not current, utilization. Third, the design is cross-sectional, so no association reported here is causal. Fourth, the pre-specified comparisons are underpowered at this sample size; accordingly, no p-value is treated as confirmatory. Fifth, acceptability was measured for hypothetical technologies; stated willingness to try such interventions may exceed real-world adoption. Sixth, the instrument was newly developed, hence its key constructs are single-item measures, and it was not cognitively tested with caregiver families, so item wording may not fully capture caregiver priorities. Finally, the sample was predominantly White (82%), recruited in English only, and universally internet-connected, and the equity literature summarized above cautions against assuming these acceptability findings transfer to the populations most affected by access gaps.

## Conclusion

In a self-selected, single-center sample of 84 caregivers of children with neurodevelopmental or behavioral conditions, respondents described frequent crises, low confidence, limited training, and substantial barriers to standard therapy, alongside high, feature-specific openness to technology-based crisis support tempered chiefly by privacy concerns.

For technology developers and behavioral-health services, these preliminary data support building in-home crisis tools around the priorities caregivers expressed, personalized plans and concrete de-escalation guidance foremost, delivered smartphone-first with optional wearable pairing and privacy protections designed in from the start. For researchers, the findings motivate larger, multi-site, demographically representative acceptability studies with registered protocols; co-design work with caregivers, including those with limited connectivity, in line with the shared-decision recommendations of qualitative work^44^; and prototype trials that pair the maturing sensing-and-coaching stack^29,30,37^ with the feature priorities reported here. These hypothesis-generating findings provide preliminary evidence of caregiver demand and a candidate starting specification for future tools and trials, one that puts personalization first and builds privacy in.

## Declarations

### Ethics approval and consent

Approved by the Cincinnati Children’s Hospital Medical Center Institutional Review Board (#2025-0646; expedited category 7). Implied consent was obtained from all participants via a study-information screen requiring an explicit “I AGREE” before survey access.

### Use of artificial intelligence

The authors used a large language model (Claude; Anthropic) under their direction to assist with analysis-code development, literature screening and citation verification, and drafting and editing of the manuscript text. Survey data were provided by human respondents and were neither generated nor altered by artificial intelligence. All statistical results were independently re-computed in a second environment (Python) and reproduced end-to-end from the raw data export, and every citation was verified against the Crossref registry. The authors reviewed, revised, and approved all content and take full responsibility for the integrity and accuracy of the work. No artificial-intelligence tool meets authorship criteria, and none is listed as an author.

### Data availability

The raw survey export contains potentially identifiable fields and cannot be shared. A de-identified derived dataset, the variable codebook documenting all derived and de-identified variables, the dated analysis plan and method-revision record, and all analysis code sufficient to reproduce the tables and figures are available from the corresponding author on reasonable request.

### Author contributions (CRediT)

Fenil Patel: Conceptualization, Methodology, Investigation, Data curation, Formal analysis, Visualization, Writing – original draft. Brittany Williams: Investigation, Writing – review & editing. Rana Elmaghraby: Investigation, Writing – review & editing. Ernest V. Pedapati: Conceptualization, Methodology, Supervision, Writing – review & editing.

## Data Availability

De-identified derived dataset and analysis code available upon reasonable request to the corresponding author.

## Acknowledgements

We thank Priya Ramesh, Clinical Research Coordinator, for her help with procurement of recruitment marketing materials, and Rebecca Patrick, LISW-S, Mark Johnson, MD, and Katherine Zappia, MD, PhD, for their help with participant recruitment.

## Supplementary Material

### Supplementary Table S1

**Table S1.**
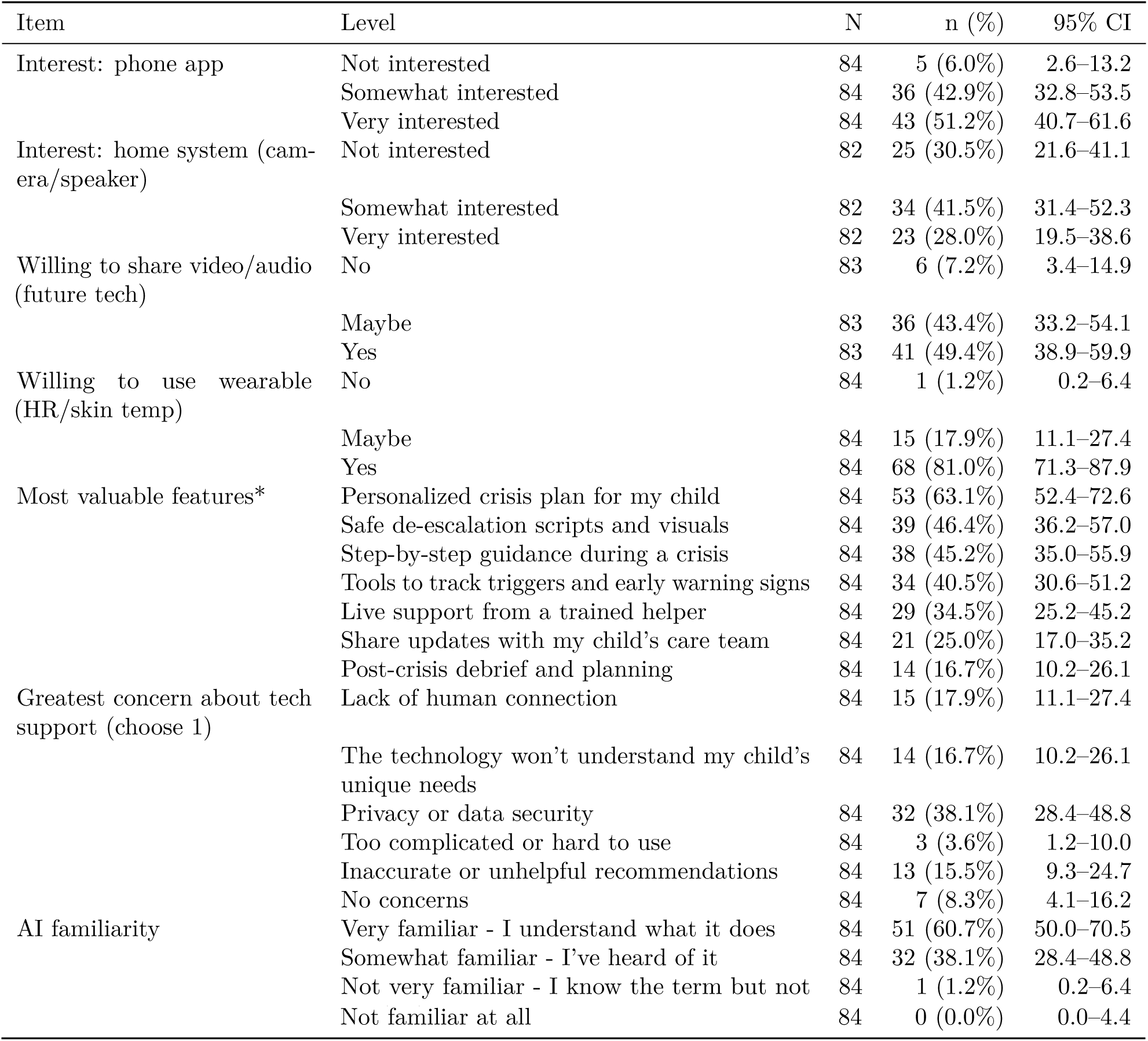
Technology acceptability detail (companion to Figures 2–3 in the main text): interest, willingness, priority features (*, up to three selections), concerns, and AI familiarity. The N column gives each item’s denominator (single-response items had 1–2 nonresponses). CI = Wilson 95% confidence interval.

### Survey instrument

The full 24-item survey instrument (Behavioral Support Survey, version 4.0, including the eligibility and consent screens) is provided as a separate supplementary file (Appendix A).

### STROBE checklist — cross-sectional studies

| # | Item | Where addressed |
| --- | --- | --- |
| 1a | Design indicated in title/abstract | Abstract Methods (“single-center cross-sectional online survey”) |
| 1b | Informative, balanced abstract | Structured abstract |
| 2 | Background/rationale | Introduction ¶1–5 |
| 3 | Objectives | Introduction, final ¶ (4 objectives) |
| 4 | Study design (early) | Methods “Study design and setting” |
| 5 | Setting, locations, dates | Methods “Study design and setting”–2.2 (CCHMC; Dec 29, 2025 – Jul 22, 2026; recruitment closed) |
| 6a | Eligibility, sources, selection | Methods “Participants and recruitment” |
| 7 | Variables defined | Methods “Variables and denominators” (incl. skip-logic denominators, derived variables) |
| 8 | Data sources / measurement | Methods “Survey instrument” (instrument, REDCap) |
| 9 | Bias | Methods “Participants and recruitment” (convenience sample; duplicate-entry safeguards; crisis history not an eligibility criterion) + “Statistical methods” transparency statement; Discussion Limitations (self-selection, self-report, recall, raffle/duplicate risk) |
| 10 | Study size | Methods “Statistical methods” (target 75 = feasibility-based enrollment target — recruitment window + clinic volume — not a power/precision calculation; closed at N = 84 caregiver responses [85 submissions – 1 study-team test entry], above target; Wilson half-width context given; underpowered, hypothesis-generating) |
| 11 | Quantitative variables handling | Methods “Variables and denominators”, “Statistical methods” (ordinal methods; medians/IQR) |
| 12a | Statistical methods | Methods “Statistical methods” |
| 12b | Subgroups/interactions | None performed (stated scope: descriptive + 3 bivariate) |
| 12c | Missing data | Methods “Data handling and missing data” (complete-case per item; N reported per estimate) |
| 12d | Sampling strategy (analytical) | N/A (no weighting; convenience sample, disclosed) |
| 12e | Sensitivity analyses | Methods “Statistical methods” (A1/A3 collapsed + Fisher); “Data handling and missing data” (partial-record exclusion: no headline proportion shifted > 0.9 pp) |
| 13 | Participants (numbers, non-response) | Results “Participants” (85 submissions; 1 study-team test entry excluded; 84 respondents, 83 complete; multi-channel recruitment, so denominator unknown and response rate not computable; screen/view counts not retained — disclosed) |
| 14 | Descriptive data | Results “Participants” + Table 1 |
| 15 | Outcome data | Results “Crisis burden and service utilization”–3.5 + Table 2, Supplementary Table S1, Figures 1–3 |
| 16a | Main results with precision (95% CI) | Wilson CIs throughout; effect sizes with CIs in Table 3 |
| 16b | Category boundaries | Instrument response options reported verbatim in tables |
| 16c | Absolute risk translation | N/A (descriptive survey) |
| 17 | Other analyses | Results “Pre-specified bivariate analyses”–3.7 (pre-specified vs exploratory clearly separated) |
| 18 | Key results summary | Discussion “Principal findings” |
| 19 | Limitations | Discussion “Limitations” (seven named limitations, incl. the unvalidated single-item instrument; duplicate-entry risk disclosed in Methods “Participants and recruitment”) |
| 20 | Cautious interpretation | Discussion “Comparison with prior work”/“Conclusion” (hypothesis-generating framing throughout) |
| 21 | Generalisability | Discussion “Limitations” (self-selected, single-center, digitally connected sample) |
| 22 | Funding | Declarations (no specific funding; REDCap grant 1UM1TR005265) |

CHERRIES checklist — internet e-surveys
| Item | Where addressed |
| --- | --- |
| Design — target population, sample frame | Methods “Participants and recruitment” (convenience sample; CCHMC neurobehavioral continuum) |
| IRB approval | Methods “Study design and setting” (CCHMC IRB #2025-0646, expedited cat. 7) |
| Informed consent | Methods “Participants and recruitment” (implied consent; study-information screen + “I AGREE”) |
| Data protection | Methods “Participants and recruitment”/“Data handling and missing data” (REDCap on CCHMC servers; raffle contacts stored separately; de-identified analysis) |
| Development & pre-testing | Methods “Survey instrument” (items developed de novo, not adapted from validated measures; single-item constructs; embedded caregiver-facing crisis definition; pre-tested by clinical colleagues for functionality and user experience; not piloted with caregiver families) |
| Open vs. closed survey | Methods “Participants and recruitment” (distributed only through clinic-based channels — flyers, email, in-person; not advertised on the open web; eligibility-screen gated) |
| Contact mode | Methods “Participants and recruitment” (flyers, email invitation, in-person clinic invitation) |
| Advertising / recruitment | Methods “Participants and recruitment” (three clinic-based channels; materials framed the study as behavioral-crisis support; raffle mentioned in materials) |
| Survey administration (web-based) | Methods “Study design and setting” (REDCap online) |
| Context | Methods “Study design and setting”–2.2 (single US pediatric center) |
| Mandatory/voluntary | Methods “Participants and recruitment” (voluntary) |
| Incentives | Methods “Participants and recruitment” (raffle, ten \$25 gift cards, completed surveys only) |
| Time frame | Methods “Study design and setting” (Dec 29, 2025 – Jul 22, 2026; recruitment closed) |
| Randomization of items | Methods “Survey instrument” (none; stated) |
| Adaptive questioning | Methods “Survey instrument” (REDCap skip logic for 2 conditional sub-items; stated) |
| Number of items | Methods “Survey instrument” (24 items; per-section counts) |
| Number of screens/pages | Methods “Survey instrument” (8 screens: eligibility, consent, 5 sections, thank-you/raffle) |
| Completeness checks | Methods “Survey instrument” (eligibility items, consent, and child age required; all other items optional) |
| Review step (back button / change answers) | Methods “Survey instrument” (back navigation and save-and-return enabled until final submission; no changes after submission) |
| Unique site visitors | Not captured; disclosed in Methods “Participants and recruitment” |
| View rate (unique visitors/invitees) | Not computable (multi-channel recruitment; denominator unknown); disclosed in Methods “Participants and recruitment” and Results “Participants” |
| Participation rate | Not computable (denominator unknown); disclosed |
| Completion rate | Results “Participants” (83/84 = 99% of caregivers who started) |
| Cookies / IP checks / duplicate prevention | Methods “Participants and recruitment” (none performed; disclosed) |
| Handling of incomplete questionnaires | Methods “Data handling and missing data” (partial record included per item; sensitivity check) |
| Questionnaires with atypical timestamp | Submission timing not captured in export; no exclusions applied |
| Statistical correction (weighting/propensity) | None used (stated; convenience sample) |

## APPENDIX A: Survey Instrument [Version 4.0_ 12/08/2025]

### Behavioral Support Survey

#### ELIGIBILITY SCREEN

1. Are you 18 years or older? [Yes/No]
2. Are you the parent/caregiver of a child (age 5 to 17 years) with behavioral or neurodevelopmental condition? [Yes/No]
3. Can you complete a survey in English? [Yes/No]

*If any “No” → “Thank you for your interest. You do not meet eligibility criteria for this study.”*

## CONSENT SCREEN

### RESEARCH STUDY INFORMATION

#### Purpose

When your child has a behavioral crisis, you need help fast. We want to know what kind of help works best. Your answers will guide us in creating tools that truly support families like yours. For this survey, a “behavioral crisis” means an episode where your child’s behavior becomes intense, unsafe, or disruptive enough that you need to act right away.

- **Time:** About 10-15 minutes
- **Compensation:** Chance to win one of ten $25 Amazon gift cards (only fully completed surveys are eligible)
- **Voluntary:** You may skip questions or stop anytime
- **Risks:** Your privacy is important. We follow health privacy rules (HIPAA: Health Insurance Portability and Accountability Act) and protect your privacy by storing all information in a secure, password-protected system. Only our research team can see your answers. As with any online data, there is a small risk of a security breach where unauthorized entities could gain access to the information.
- **Benefits:** Your responses may help develop better crisis support tools for families in the future.
- **Contact:**
- **IRB:** Approved by CCHMC IRB, Study # 2025-0646

**Participation is voluntary. You must be the parent/caregiver of a child (age 5 to 17 years) with behavioral or neurodevelopmental conditions to participate. You may skip any question or stop at any time without affecting your child’s care at Cincinnati Children’s.**

**Your responses will be stored in a secure, HIPAA-compliant database. Only the research team will have access to identified data. Your consent to participate in this survey serves as authorization to collect health information.**

By continuing, you consent to participate. [I AGREE] [I DO NOT AGREE]

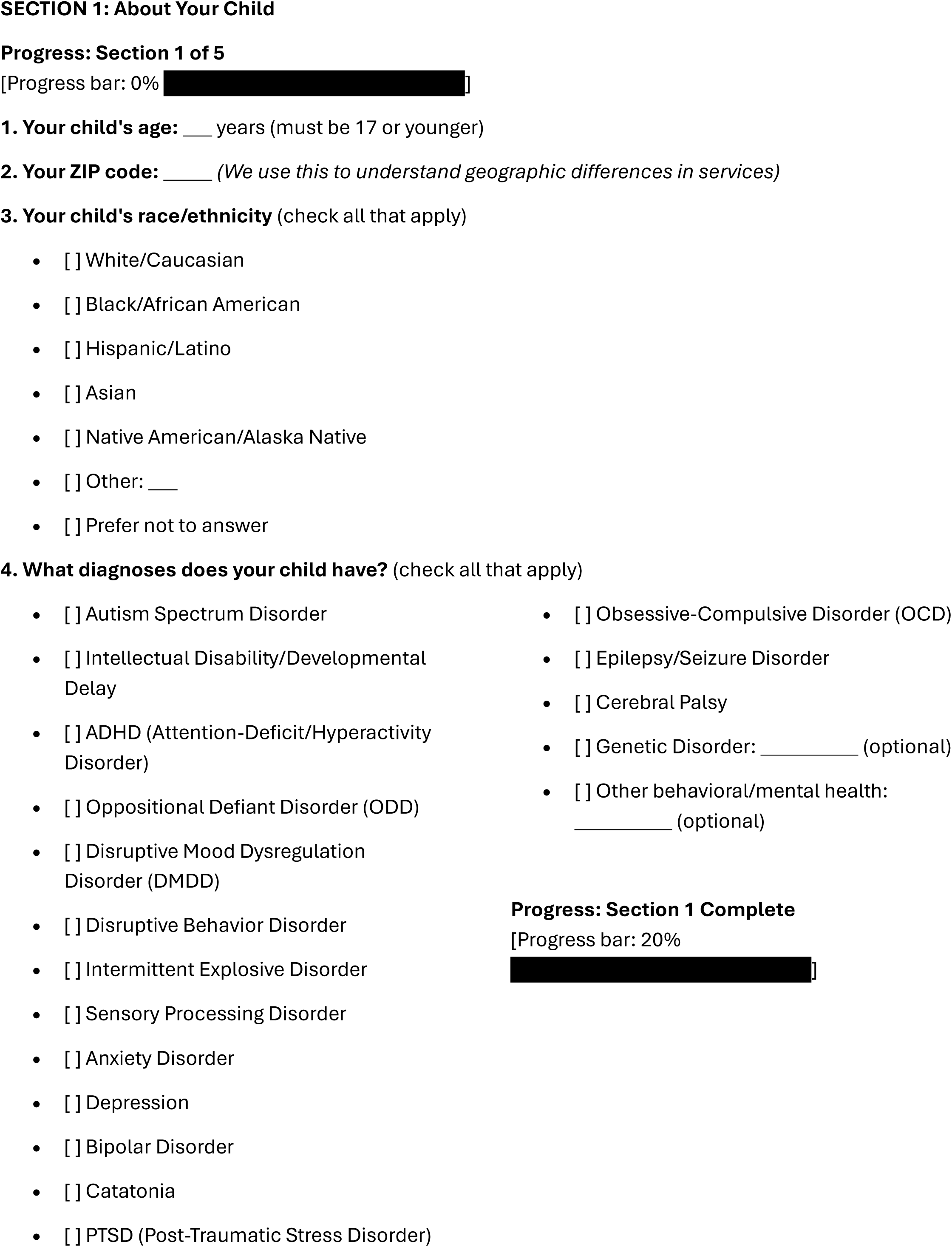

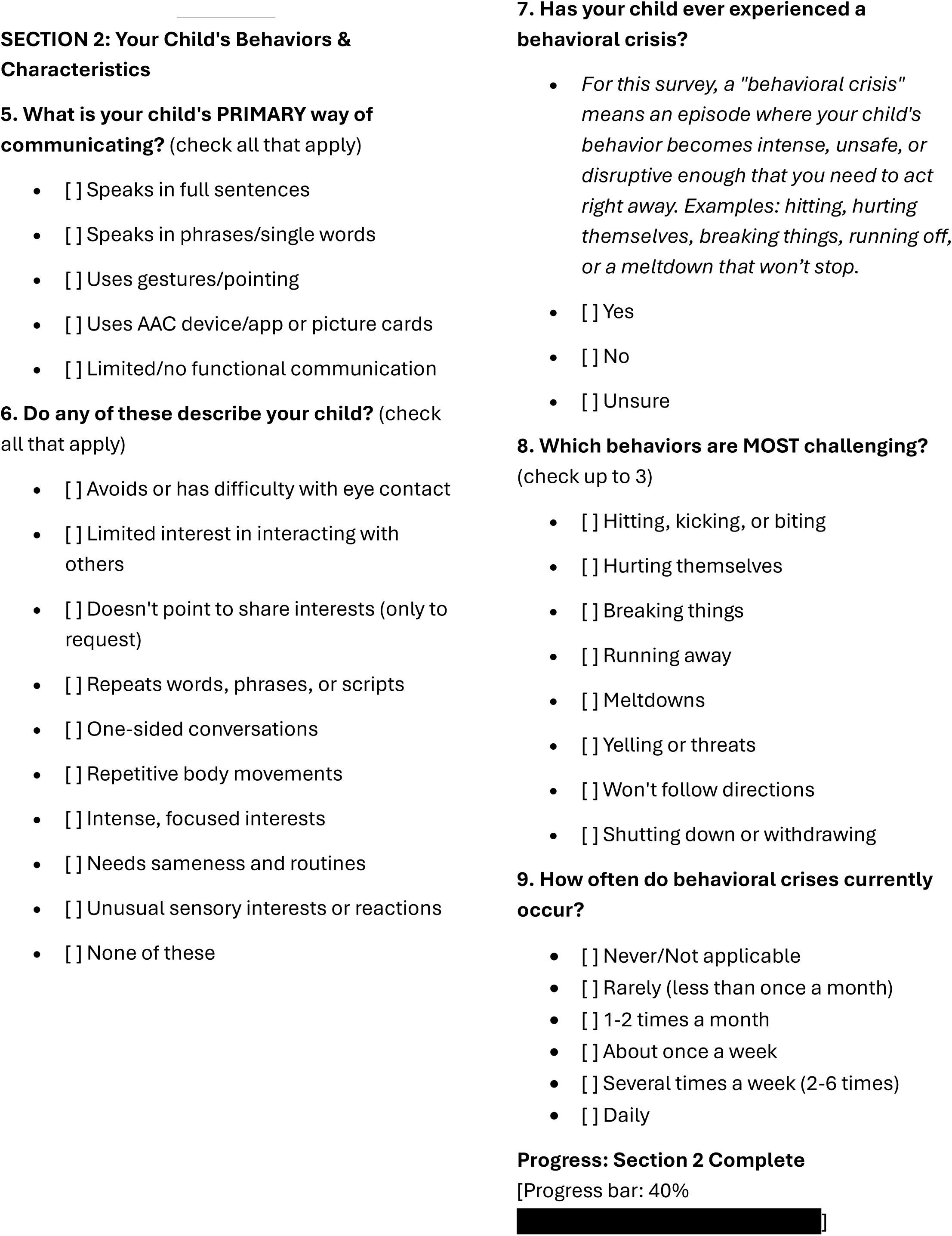

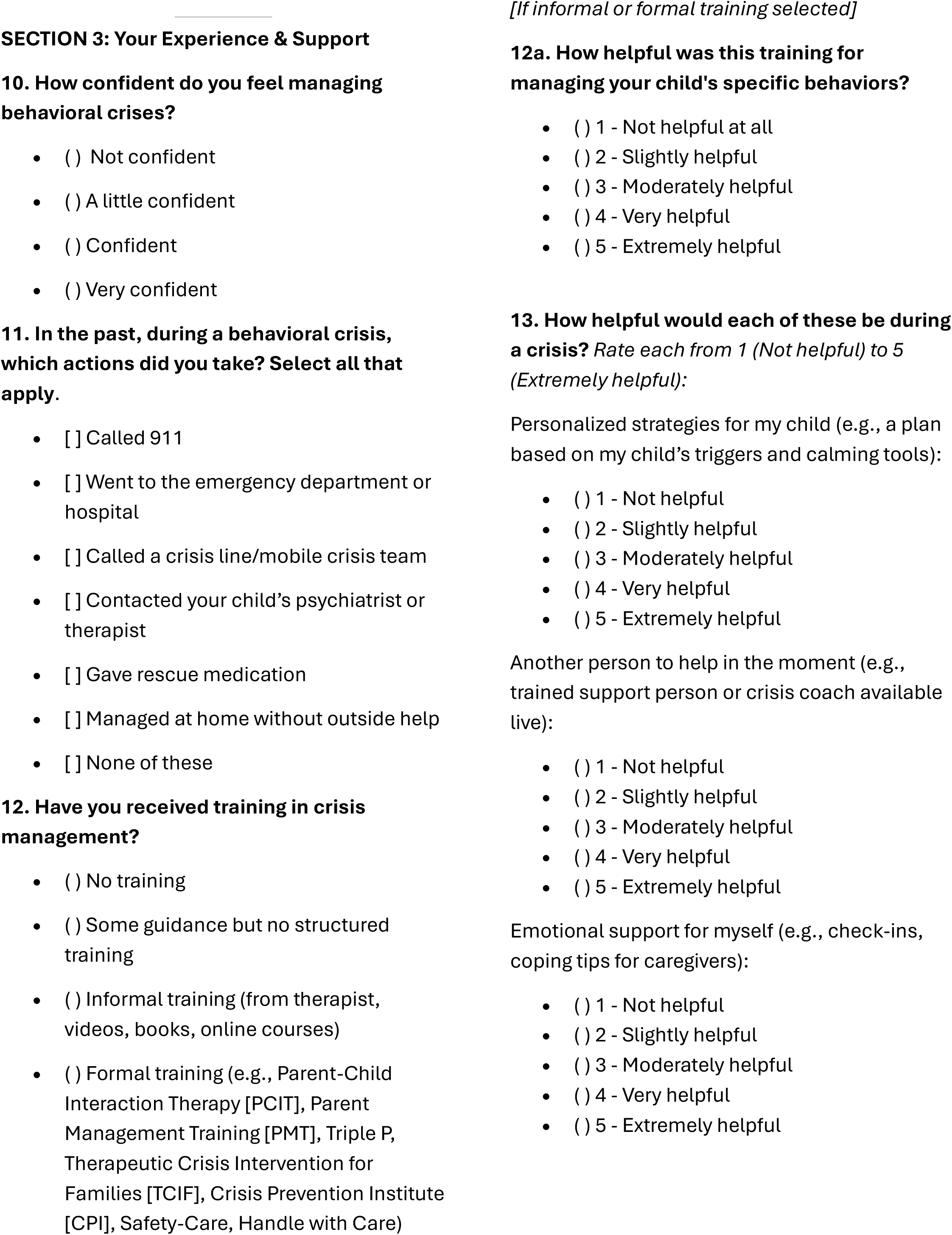

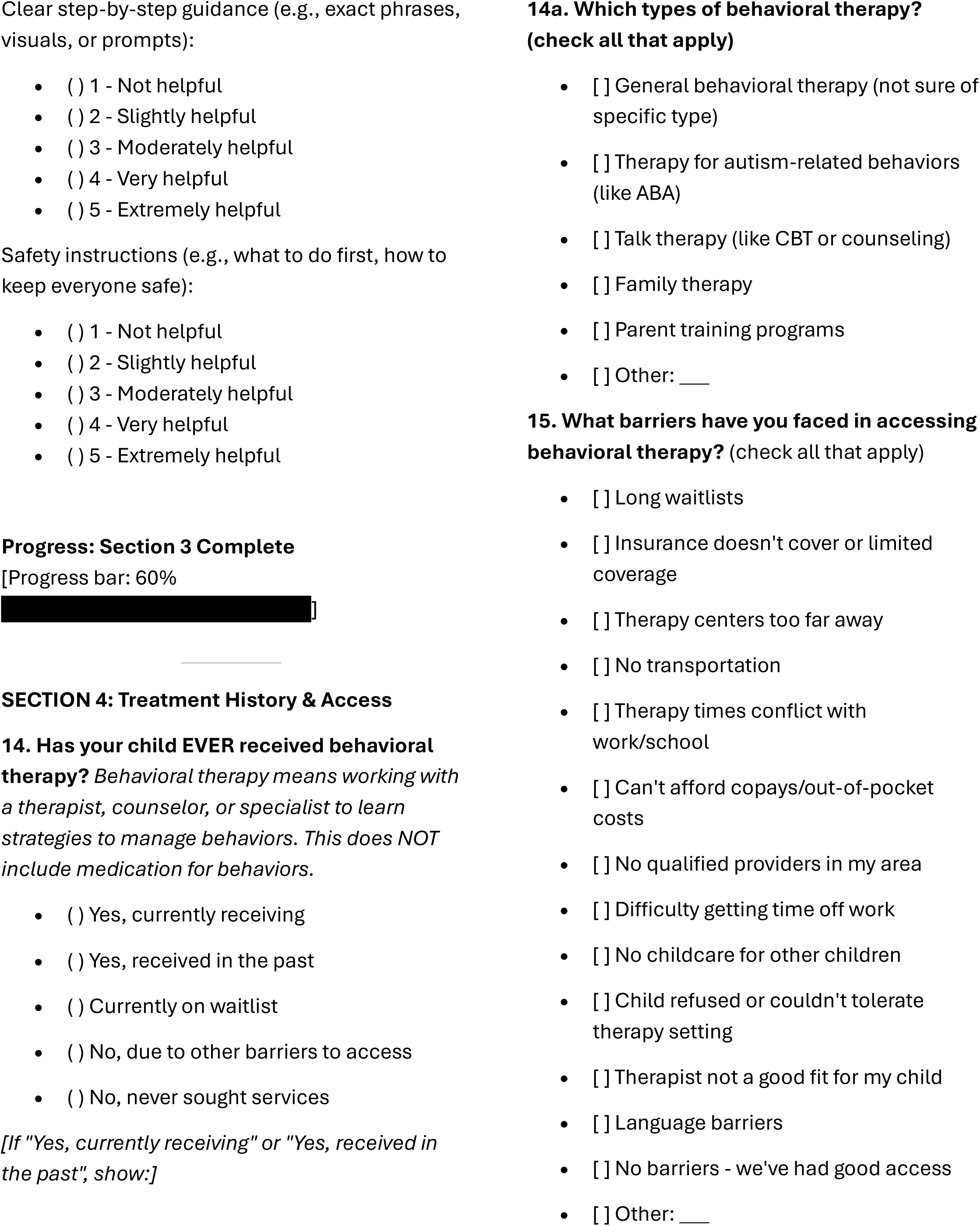

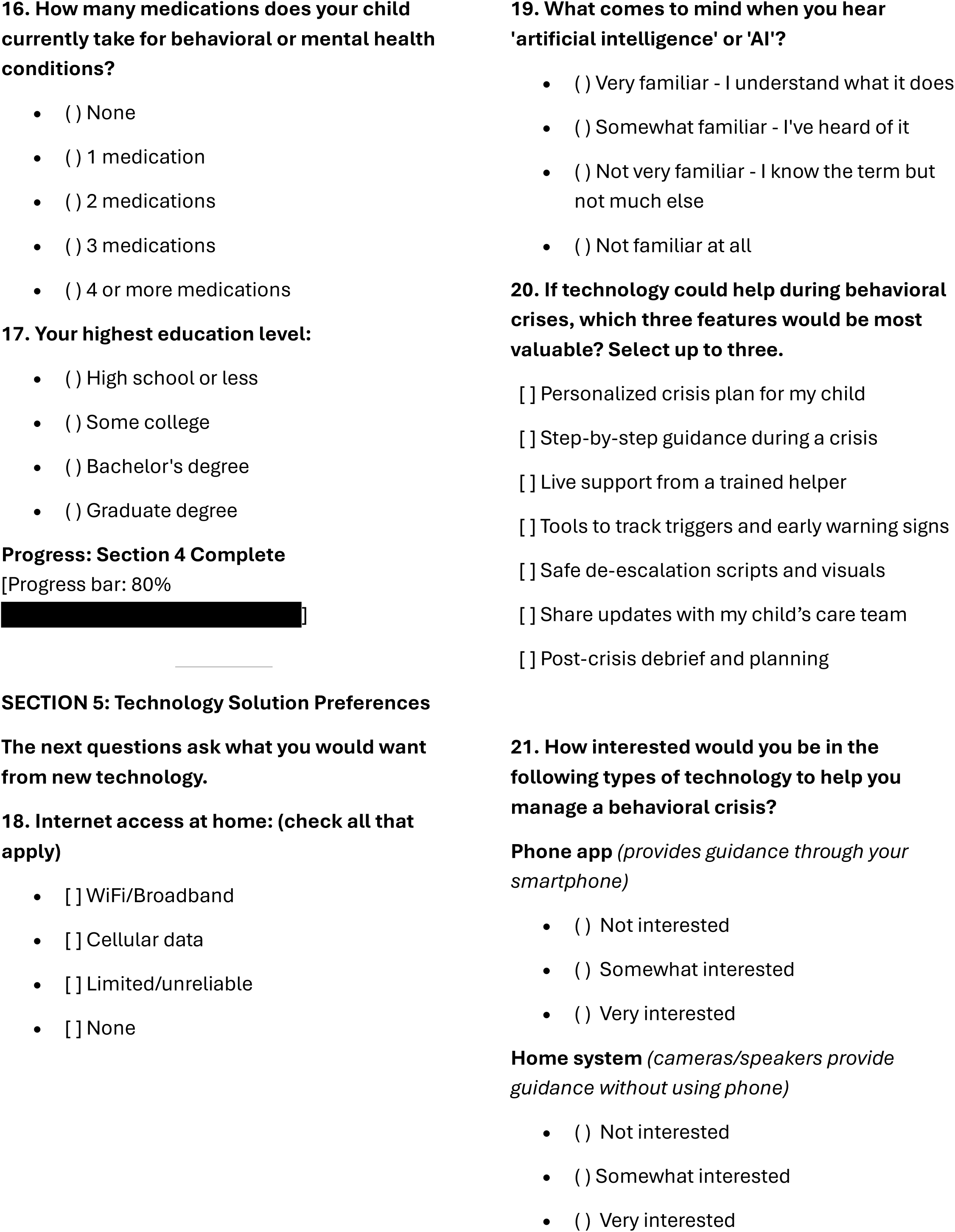

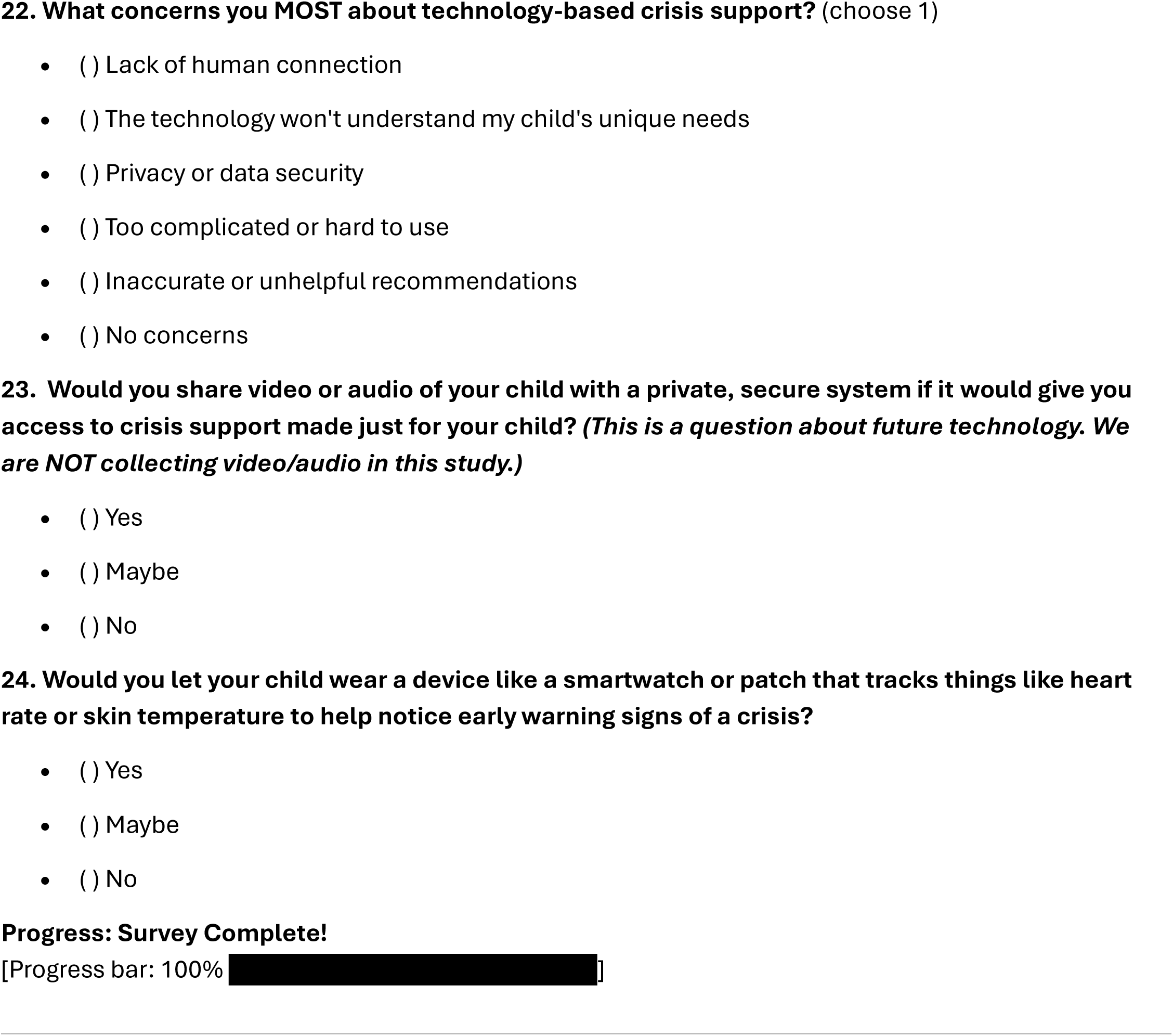

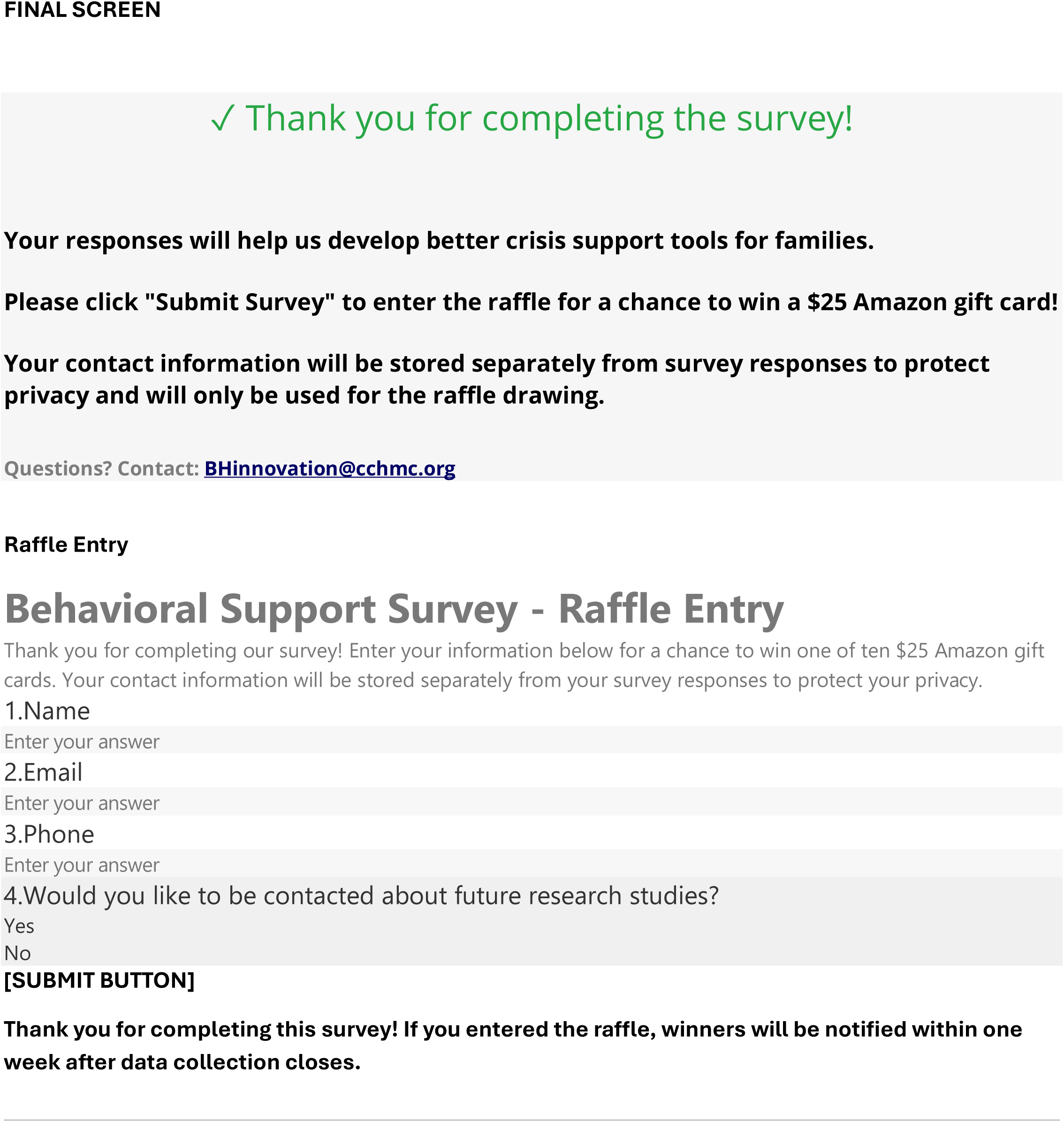

## References

1. Catherine Steenfeldt-Kristensen, Chris A. Jones, Caroline Richards. The prevalence of self-injurious behaviour in autism: A meta-analytic study. Journal of Autism and Developmental Disorders. 2020;50(11):3857–3873. doi:10.1007/s10803-020-04443-1

2. Stephen M. Kanne, Micah O. Mazurek. Aggression in children and adolescents with ASD: Prevalence and risk factors. Journal of Autism and Developmental Disorders. 2010;41(7):926–937. doi:10.1007/s10803-010-1118-4

3. Bridget Kiely, Talia R. Migdal, Sujit Vettam, Andrew Adesman. Prevalence and correlates of elopement in a nationally representative sample of children with developmental disabilities in the united states, ed.Andrea Martinuzzi. PLOS ONE. 2016;11(2):e0148337. doi:10.1371/journal.pone.0148337

4. Laura McLaughlin, Sarah A. Keim, Andrew Adesman. Wandering by children with autism spectrum disorder: Key clinical factors and the role of schools and pediatricians. Journal of Developmental & Behavioral Pediatrics. 2018;39(7):538–546. doi:10.1097/dbp.0000000000000591

5. Roma A. Vasa, Louis Hagopian, Luke G. Kalb. Investigating mental health crisis in youth with autism spectrum disorder. Autism Research. 2019;13(1):112–121. doi:10.1002/aur.2224

6. Luther G. Kalb, Elizabeth A. Stuart, Brian Freedman, Benjamin Zablotsky, Roma Vasa. Psychiatric-related emergency department visits among children with an autism spectrum disorder. Pediatric Emergency Care. 2012;28(12):1269–1276. doi:10.1097/pec.0b013e3182767d96

7. Scott Lindgren, Emily Lauer, Elizabeth Momany, Tara Cope, Julie Royer, Lindsay Cogan, Suzanne McDermott, Brian S. Armour. Disability, hospital care, and cost: Utilization of emergency and inpatient care by a cohort of children with intellectual and developmental disabilities. The Journal of Pediatrics. 2021;229:259–266. doi:10.1016/j.jpeds.2020.08.084

8. Victor Hong, Fiona Miller, Shane Kentopp, Hannah Reynard, Bernard Biermann, Can Beser, Saad Shamshair, Bailey Fay, Ahmad Shobassy, Michelle Stanley, Cody Weston, Mohammad Ghaziuddin, Neera Ghaziuddin. Patients with autism spectrum or intellectual disability in the psychiatric emergency department: Findings from a 10-year retrospective review. Journal of Autism and Developmental Disorders. 2024;56(5):1892–1901. doi:10.1007/s10803-024-06658-y

9. Paul Turcotte, Lindsay L. Shea, David Mandell. School discipline, hospitalization, and police contact overlap among individuals with autism spectrum disorder. Journal of Autism and Developmental Disorders. 2017;48(3):883–891. doi:10.1007/s10803-017-3359-y

10. Karen Bearss, Cynthia Johnson, Tristram Smith, Luc Lecavalier, Naomi Swiezy, Michael Aman, David B. McAdam, Eric Butter, Charmaine Stillitano, Noha Minshawi, Denis G. Sukhodolsky, Daniel W. Mruzek, Kylan Turner, Tiffany Neal, Victoria Hallett, James A. Mulick, Bryson Green, Benjamin Handen, Yanhong Deng, James Dziura, Lawrence Scahill. Effect of parent training vs parent education on behavioral problems in children with autism spectrum disorder: A randomized clinical trial. JAMA. 2015;313(15):1524. doi:10.1001/jama.2015.3150

11. Korrie Allen, John Harrington, Lauren B. Quetsch, Joshua Masse, Cathy Cooke, James F. Paulson. Parent–child interaction therapy for children with disruptive behaviors and autism: A randomized clinical trial. Journal of Autism and Developmental Disorders. 2022;53(1):390–404. doi:10.1007/s10803-022-05428-y

12. Simone Breider, Annelies de Bildt, Kirstin Greaves-Lord, Andrea Dietrich, Pieter J. Hoekstra, Barbara J. van den Hoofdakker. Parent training for disruptive behaviors in referred children with autism spectrum disorder: A randomized controlled trial. Journal of Autism and Developmental Disorders. 2024;56(2):481–498. doi:10.1007/s10803-024-06567-0

13. Marissa E Yingling, Matthew H Ruther, Erick M Dubuque, David S Mandell. County-level variation in geographic access to board certified behavior analysts among children with autism spectrum disorder in the united states. Autism. 2021;25(6):1734–1745. doi:10.1177/13623613211002051

14. Marissa E. Yingling, Matthew H. Ruther, Erick M. Dubuque. Trends in geographic access to board certified behavior analysts among children with autism spectrum disorder, 2018–2021. Journal of Autism and Developmental Disorders. 2022;52(12):5483–5490. doi:10.1007/s10803-021-05402-0

15. Ryan K. McBain, Jonathan H. Cantor, Aaron Kofner, Bradley D. Stein, Hao Yu. State insurance mandates and the workforce for children with autism. Pediatrics. 2020;146(4). doi:10.1542/peds.2020-0836

16. Wanqing Zhang, Grace Baranek. The impact of insurance coverage types on access to and utilization of health services for u.s. Children with autism. Psychiatric Services. 2016;67(8):908–911. doi:10.1176/appi.ps.201500206

17. Teal W. Benevides, Henry J. Carretta, Shelly J. Lane. Unmet need for therapy among children with autism spectrum disorder: Results from the 2005–2006 and 2009–2010 national survey of children with special health care needs. Maternal and Child Health Journal. 2015;20(4):878–888. doi:10.1007/s10995-015-1876-x

18. Justine Brennan, Olivia F. Ward, Theodore S. Tomeny, Thompson E. Davis. A systematic review of parental self-efficacy in parents of autistic children. Clinical Child and Family Psychology Review. 2024;27(3):878–905. doi:10.1007/s10567-024-00495-2

19. Thumar Bhavika Jivanlal, Jharna Ganguly. Mental health challenges faced by caregivers of children with intellectual disability, ADHD, and autism. Vidhyayana. 2026;11(si2). doi:10.58213/nydeaq96

20. Ameer S. J. Hohlfeld, Michal Harty, Mark E. Engel. Parents of children with disabilities: A systematic review of parenting interventions and self-efficacy. African Journal of Disability. 2018;7. doi:10.4102/ajod.v7i0.437

21. Olivia J Lindly, Amy M Shui, Noa M Stotts, Karen A Kuhlthau. Caregiver strain among north american parents of children from the autism treatment network registry call-back study. Autism. 2021;26(6):1460–1476. doi:10.1177/13623613211052108

22. Tamara Ondruskova, Rachel Royston, Michael Absoud, Gareth Ambler, Chen Qu, Jacqueline Barnes, Rachael Hunter, Monica Panca, Marinos Kyriakopoulos, Kate Oulton, Eleni Paliokosta, Aditya Narain Sharma, Vicky Slonims, Una Summerson, Alastair Sutcliffe, Megan Thomas, Brindha Dhandapani, Helen Leonard, Angela Hassiotis. Clinical and cost-effectiveness of an adapted intervention for preschoolers with moderate to severe intellectual disabilities displaying behaviours that challenge: The EPICC-ID RCT. Health Technology Assessment. Published online January 2024:1–94. doi:10.3310/jkty6144

23. Angela V. Dahiya, Rosanna Breaux, Stephanie N. Pham, Daniele C. Martino, Megan Fok, Jordan Albright, Delshad M. Shroff, Angela Scarpa. Using a mobile app to support parents of children with behavior problems. Research on Child and Adolescent Psychopathology. 2025;53(12):1879–1892. doi:10.1007/s10802-025-01385-z

24. Stéphanie Turgeon, Marc J. Lanovaz, Marie-Michèle Dufour. Effects of an interactive web training to support parents in reducing challenging behaviors in children with autism. Behavior Modification. 2020;45(5):769–796. doi:10.1177/0145445520915671

25. JooHyun Lee, JaeHyun Lim, Soyeon Kang, Sujin Kim, So Yoon Jung, Sujin Kim, Soon-Beom Hong, Yu Rang Park. Mobile app–assisted parent training intervention for behavioral problems in children with autism spectrum disorder: Pilot randomized controlled trial. JMIR Human Factors. 2024;11:e52295. doi:10.2196/52295

26. Rinat Meerson, Hanna Buchholz, Klaus Kammerer, Manuel Göster, Johannes Schobel, Christoph Ratz, Rüdiger Pryss, Regina Taurines, Marcel Romanos, Matthias Gamer, Julia Geissler. ProVIA-kids - outcomes of an uncontrolled study on smartphone-based behaviour analysis for challenging behaviour in children with intellectual and developmental disabilities or autism spectrum disorder. Frontiers in Digital Health. 2024;6. doi:10.3389/fdgth.2024.1462682

27. Eunkyung Jo, Seora Park, Hyeonseok Bang, Youngeun Hong, Yeni Kim, Jungwon Choi, Bung Nyun Kim, Daniel A. Epstein, Hwajung Hong. GeniAuti: Toward data-driven interventions to challenging behaviors of autistic children through caregivers’ tracking. Proceedings of the ACM on Human-Computer Interaction. 2022;6(CSCW1):1–27. doi:10.1145/3512939

28. Inbal Nahum-Shani, Shawna N Smith, Bonnie J Spring, Linda M Collins, Katie Witkiewitz, Ambuj Tewari, Susan A Murphy. Just-in-time adaptive interventions (JITAIs) in mobile health: Key components and design principles for ongoing health behavior support. Annals of Behavioral Medicine. 2017;52(6):446–462. doi:10.1007/s12160-016-9830-8

29. Inbal Nahum-Shani, Susan A. Murphy. Just-in-time adaptive interventions: Where are we now and what is next? Annual Review of Psychology. 2026;77(1):679–703. doi:10.1146/annurev-psych-121024-044244

30. Magdalena Romanowicz, Maria T. Saliba, Angelina R. Wilton, Juan F. Garzon Hincapie, Kyle S. Croarkin, Christina T. Saliba, Allison LeMahieu, Noelle Drapeau, Brandi Schlichting, Michelle Skime, William V. Bobo, Jennifer L. Vande Voort, Julia Shekunov, Paul E. Croarkin, Arjun P. Athreya. Feasibility of digital augmentation of parent-child interaction therapy: A randomized clinical trial. JAMA Network Open. 2025;8(12):e2548869. doi:10.1001/jamanetworkopen.2025.48869

31. Md Mehedi Hassan, Md Nayeem Hasan, Sanjida Islam, Abdullah Hill Hussain, Md Maruful Islam. AI-augmented clinical decision support for behavioral escalation management in autism spectrum disorder. Journal of International Crisis and Risk Communication Research. Published online December 2023:201–208. doi:10.63278/jicrcr.vi.3312

32. Zhaobo K. Zheng, John E. Staubitz, Amy S. Weitlauf, Johanna Staubitz, Marney Pollack, Lauren Shibley, Michelle Hopton, William Martin, Amy Swanson, Pablo Juárez, Zachary E. Warren, Nilanjan Sarkar. A predictive multimodal framework to alert caregivers of problem behaviors for children with ASD (PreMAC). Sensors. 2021;21(2):370. doi:10.3390/s21020370

33. Shadi Ghafghazi, Amarie Carnett, Leslie Neely, Arun Das, Paul Rad. AI-augmented behavior analysis for children with developmental disabilities: Building toward precision treatment. *IEEE Systems*, Man, and Cybernetics Magazine. 2021;7(4):4–12. doi:10.1109/msmc.2021.3086989

34. Freddy Jackson Brown, Isabelle Stewart Muscat, Louise Quinn, Lara Best, Paul Cooper. Validation of the safety, empathy and utility of a large language model conversation agent for parents of autistic and neurodivergent children. Scientific Reports. 2026;16(1). doi:10.1038/s41598-026-44254-5

35. Matthew S. Goodwin, Carla A. Mazefsky, Stratis Ioannidis, Deniz Erdogmus, Matthew Siegel. Predicting aggression to others in youth with autism using a wearable biosensor. Autism Research. 2019;12(8):1286–1296. doi:10.1002/aur.2151

36. Tales Imbiriba, Ahmet Demirkaya, Ashutosh Singh, Deniz Erdogmus, Matthew S. Goodwin. Wearable biosensing to predict imminent aggressive behavior in psychiatric inpatient youths with autism. JAMA Network Open. 2023;6(12):e2348898. doi:10.1001/jamanetworkopen.2023.48898

37. Nibraas Khan, Abigale Plunk, Zhaobo Zheng, Deeksha Adiani, John Staubitz, Amy Weitlauf, Nilanjan Sarkar. Pilot study of a real-time early agitation capture technology (REACT) for children with intellectual and developmental disabilities. DIGITAL HEALTH. 2024;10. doi:10.1177/20552076241287884

38. Moid Sandhu, Siddique Latif, Andrew Bayor, Wei Lu, Mahnoosh Kholghi, Deepa Prabhu, David Silvera-Tawil. Empowering caregivers of individuals with autism spectrum disorder through sensor-based monitoring of emotional dysregulation: A scoping review. International Journal of Medical Informatics. 2026;209:106262. doi:10.1016/j.ijmedinf.2026.106262

39. Patrick W. Romani, Sidney K. D’Mello, Robert M. Moulder, Lily N. Berkowitz. Using wearable technology to predict the occurrence of severe behavior problems among neurodiverse individuals: A systematic review. Perspectives on Behavior Science. 2026;49(2):361–383. doi:10.1007/s40614-026-00497-1

40. Paul A. Harris, Robert Taylor, Robert Thielke, Jonathon Payne, Nathaniel Gonzalez, Jose G. Conde. Research electronic data capture (REDCap)—a metadata-driven methodology and workflow process for providing translational research informatics support. Journal of Biomedical Informatics. 2009;42(2):377–381. doi:10.1016/j.jbi.2008.08.010

41. Erik von Elm, Douglas G. Altman, Matthias Egger, Stuart J. Pocock, Peter C. Gøtzsche, Jan P. and Vandenbroucke. The strengthening the reporting of observational studies in epidemiology (STROBE) statement: Guidelines for reporting observational studies. Annals of Internal Medicine. 2007;147(8):573–577. doi:10.7326/0003-4819-147-8-200710160-00010

42. Gunther Eysenbach. Improving the quality of web surveys: The checklist for reporting results of internet e-surveys (CHERRIES). Journal of Medical Internet Research. 2004;6(3):e34. doi:10.2196/jmir.6.3.e34

43. Tarek Turk, Mohamed Tamer Elhady, Sherwet Rashed, Mariam Abdelkhalek, Somia Ahmed Nasef, Ashraf Mohamed Khallaf, Abdelrahman Tarek Mohammed, Andrew Wassef Attia, Purushottam Adhikari, Mohamed Alsabbahi Amin, Kenji Hirayama, Nguyen Tien Huy. Quality of reporting web-based and non-web-based survey studies: What authors, reviewers and consumers should consider, ed.G. J. Melendez-Torres. PLOS ONE. 2018;13(6):e0194239. doi:10.1371/journal.pone.0194239

44. Rachel Xifaras, David J. Amor, Erin Turbitt, Claudine M. Kraan. Parents’ experiences and views about use of wearable technology for research and treatment monitoring of children with neurodevelopmental disorders. Journal of Developmental & Behavioral Pediatrics. 2025;46(1):e4–e9. doi:10.1097/dbp.0000000000001337

45. Hannah I. Levin, Dominique Egger, Lara Andres, Mckensey Johnson, Sarah Kate Bearman, Kaya de Barbaro. Sensing everyday activity: Parent perceptions and feasibility. Infant Behavior and Development. 2021;62:101511. doi:10.1016/j.infbeh.2020.101511

46. Laurel A. Fish, Emily J. H. Jones. A survey on the attitudes of parents with young children on in-home monitoring technologies and study designs for infant research, ed.Barbara Schouten. PLOS ONE. 2021;16(2):e0245793. doi:10.1371/journal.pone.0245793

47. Gabriella B. Smith, Mickayla D. Jones, Mary J. Akel, Leonardo Barrera, Marie Heffernan, Patrick Seed, Michelle L. Macy, Stephanie A. Fisher, Leena B. Mithal. Parental perceptions of early childhood in-home research with monitoring: A qualitative study. The Journal of Pediatrics. 2025;278:114437. doi:10.1016/j.jpeds.2024.114437

48. Esmee Adam, Franka Meiland, Noud Frielink, Erwin Meinders, Reon Smits, Petri Embregts, Hanneke Smaling. User requirements and perceptions of a sensor system for early stress detection in people with dementia and people with intellectual disability: Qualitative study. JMIR Formative Research. 2024;8:e52248. doi:10.2196/52248

49. Emily Wu, John Torous, Rashad Hardaway, Thomas Gutheil. Confidentiality and privacy for smartphone applications in child and adolescent psychiatry. Child and Adolescent Psychiatric Clinics of North America. 2017;26(1):117–124. doi:10.1016/j.chc.2016.07.006

50. Scott S. Hall, Katerina D. Monlux, Arlette Bujanda Rodriguez, Booil Jo, Joy S. Pollard. Telehealth-enabled behavioral treatment for problem behaviors in boys with fragile x syndrome: A randomized controlled trial. Journal of Neurodevelopmental Disorders. 2020;12(1). doi:10.1186/s11689-020-09331-4

51. Scott Lindgren, David Wacker, Alyssa Suess, Kelly Schieltz, Kelly Pelzel, Todd Kopelman, John Lee, Patrick Romani, Debra Waldron. Telehealth and autism: Treating challenging behavior at lower cost. Pediatrics. 2016;137(Supplement_2):S167-S175. doi:10.1542/peds.2015-2851o

52. Kait Gould, Ryan J. Martin, Summer Bottini, Jaime Crowley-Zalaket, Ainsley Losh, Meka McCammon, Jennifer R. Wolgemuth, Cynthia Anderson. Behavioral parent training via telehealth for autistic children in rural appalachia: A mixed methods feasibility study. Journal of Positive Behavior Interventions. 2023;26(4):204–217. doi:10.1177/10983007231200541

53. Glory Okwori, Sylvester Orimaye. Telehealth utilization among children with mental/behavioral health disorders. Telemedicine and e-Health. 2025;31(11):1297–1308. doi:10.1177/15305627251361297

54. Chizimuzo T. C. Okoli, Tianyi Wang, Bassema Abufarsakh, Sarret Seng, Zainab S. Almogheer, Jarrah Al-Kayed, Pooja Bhattarai, Holly Stith, Andrew Makowski. Changes in telehealth usage among medicaid beneficiaries with mental illnesses in a rural state: An analysis of kentucky medicaid dataset over 5 years (2018–2022). The Journal of Rural Health. 2025;42(1). doi:10.1111/jrh.70108

55. Bennett M. Liu, Kelley Paskov, Jack Kent, Maya McNealis, Soren Sutaria, Olivia Dods, Christopher Harjadi, Nate Stockham, Andrey Ostrovsky, Dennis P. Wall. Racial and ethnic disparities in geographic access to autism resources across the US. JAMA Network Open. 2023;6(1):e2251182. doi:10.1001/jamanetworkopen.2022.51182

56. Marissa E. Yingling, Matthew H. Ruther, Erick M. Dubuque, Bethany A. Bell. Impact of county sociodemographic factors and state policy on geographic access to behavior analysts among children with autism spectrum disorder. Administration and Policy in Mental Health and Mental Health Services Research. 2021;48(6):1105–1114. doi:10.1007/s10488-021-01120-y

57. Lucy A. Bilaver, Sarah A. Sobotka, David S. Mandell. Understanding racial and ethnic disparities in autism-related service use among medicaid-enrolled children. Journal of Autism and Developmental Disorders. 2020;51(9):3341–3355. doi:10.1007/s10803-020-04797-6

58. Kathryn A. Smith, Jean-G. Gehricke, Suzannah Iadarola, Audrey Wolfe, Karen A. Kuhlthau. Disparities in service use among children with autism: A systematic review. Pediatrics. 2020;145(Supplement_1):S35-S46. doi:10.1542/peds.2019-1895g

59. Ryan K. McBain, Jonathan H. Cantor, Aaron Kofner, Bradley D. Stein, Hao Yu. Ongoing disparities in digital and in-person access to child psychiatric services in the united states. Journal of the American Academy of Child & Adolescent Psychiatry. 2022;61(7):926–933. doi:10.1016/j.jaac.2021.11.028

60. Michelle LaClair, David S. Mandell, Andrew W. Dick, Khaled Iskandarani, Bradley D. Stein, Douglas L. Leslie. The effect of medicaid waivers on ameliorating racial/ethnic disparities among children with autism. Health Services Research. 2019;54(4):912–919. doi:10.1111/1475-6773.13176

