## Appendix A: Survey instrument (Behavioral Support Survey) for "Caregiver Needs and Technology Acceptability for Behavioral-Crisis Support in Children With Neurodevelopmental or Behavioral Conditions" (from another version of this preprint: version 1, 2026-07-13)

**APPENDIX A: Survey Instrument [Version 4.0_ 12/08/2025]**

**Behavioral Support Survey**

**ELIGIBILITY SCREEN**

1. Are you 18 years or older? [Yes/No]
2. Are you the parent/caregiver of a child (age 5 to 17 years) with behavioral or neurodevelopmental condition? [Yes/No]
3. Can you complete a survey in English? [Yes/No]

*If any "No" → "Thank you for your interest. You do not meet eligibility criteria for this study."*

---

**CONSENT SCREEN**

**RESEARCH STUDY INFORMATION**

**Purpose:** When your child has a behavioral crisis, you need help fast. We want to know what kind of help works best. Your answers will guide us in creating tools that truly support families like yours. For this survey, a "behavioral crisis" means an episode where your child's behavior becomes intense, unsafe, or disruptive enough that you need to act right away.

- **Time:** About 10-15 minutes
- **Compensation:** Chance to win one of ten $25 Amazon gift cards (only fully completed surveys are eligible)
- **Voluntary:** You may skip questions or stop anytime
- **Risks:** Your privacy is important. We follow health privacy rules (HIPAA: Health Insurance Portability and Accountability Act) and protect your privacy by storing all information in a secure, password-protected system. Only our research team can see your answers. As with any online data, there is a small risk of a security breach where unauthorized entities could gain access to the information.
- **Benefits:** Your responses may help develop better crisis support tools for families in the future.
- **Contact:**
- **IRB:** Approved by CCHMC IRB, Study # 2025-0646

**Participation is voluntary. You must be the parent/caregiver of a child (age 5 to 17 years) with behavioral or neurodevelopmental conditions to participate. You may skip any question or stop at any time without affecting your child's care at Cincinnati Children's.**

**Your responses will be stored in a secure, HIPAA-compliant database. Only the research team will have access to identified data. Your consent to participate in this survey serves as authorization to collect health information.**

By continuing, you consent to participate. [I AGREE] [I DO NOT AGREE]

#### SECTION 1: About Your Child

**Progress: Section 1 of 5**
[Progress bar: 0% [redacted]]

**1. Your child's age:** ___ years (must be 17 or younger)

**2. Your ZIP code:** _____ *(We use this to understand geographic differences in services)*

**3. Your child's race/ethnicity** (check all that apply)

- [ ] White/Caucasian
- [ ] Black/African American
- [ ] Hispanic/Latino
- [ ] Asian
- [ ] Native American/Alaska Native
- [ ] Other: ___
- [ ] Prefer not to answer

**4. What diagnoses does your child have?** (check all that apply)

- [ ] Autism Spectrum Disorder
- [ ] Intellectual Disability/Developmental Delay
- [ ] ADHD (Attention-Deficit/Hyperactivity Disorder)
- [ ] Oppositional Defiant Disorder (ODD)
- [ ] Disruptive Mood Dysregulation Disorder (DMDD)
- [ ] Disruptive Behavior Disorder
- [ ] Intermittent Explosive Disorder
- [ ] Sensory Processing Disorder
- [ ] Anxiety Disorder
- [ ] Depression
- [ ] Bipolar Disorder
- [ ] Catatonia
- [ ] PTSD (Post-Traumatic Stress Disorder)
- [ ] Obsessive-Compulsive Disorder (OCD)
- [ ] Epilepsy/Seizure Disorder
- [ ] Cerebral Palsy
- [ ] Genetic Disorder: __________ (optional)
- [ ] Other behavioral/mental health: __________ (optional)

**Progress: Section 1 Complete**
[Progress bar: 20% [redacted]]

#### SECTION 2: Your Child's Behaviors & Characteristics

**5. What is your child's PRIMARY way of communicating?** (check all that apply)

- [ ] Speaks in full sentences
- [ ] Speaks in phrases/single words
- [ ] Uses gestures/pointing
- [ ] Uses AAC device/app or picture cards
- [ ] Limited/no functional communication

**6. Do any of these describe your child?** (check all that apply)

- [ ] Avoids or has difficulty with eye contact
- [ ] Limited interest in interacting with others
- [ ] Doesn't point to share interests (only to request)
- [ ] Repeats words, phrases, or scripts
- [ ] One-sided conversations
- [ ] Repetitive body movements
- [ ] Intense, focused interests
- [ ] Needs sameness and routines
- [ ] Unusual sensory interests or reactions
- [ ] None of these

**7. Has your child ever experienced a behavioral crisis?**

- *For this survey, a "behavioral crisis" means an episode where your child's behavior becomes intense, unsafe, or disruptive enough that you need to act right away. Examples: hitting, hurting themselves, breaking things, running off, or a meltdown that won't stop.*
- [ ] Yes
- [ ] No
- [ ] Unsure

**8. Which behaviors are MOST challenging?** (check up to 3)

- [ ] Hitting, kicking, or biting
- [ ] Hurting themselves
- [ ] Breaking things
- [ ] Running away
- [ ] Meltdowns
- [ ] Yelling or threats
- [ ] Won't follow directions
- [ ] Shutting down or withdrawing

**9. How often do behavioral crises currently occur?**

- [ ] Never/Not applicable
- [ ] Rarely (less than once a month)
- [ ] 1-2 times a month
- [ ] About once a week
- [ ] Several times a week (2-6 times)
- [ ] Daily

**Progress: Section 2 Complete**
[Progress bar: 40% [redacted]]

**SECTION 3: Your Experience & Support**

**10. How confident do you feel managing behavioral crises?**

- ( )  Not confident
- ( ) A little confident
- ( ) Confident
- ( ) Very confident

**11. In the past, during a behavioral crisis, which actions did you take? Select all that apply**.

- [ ] Called 911
- [ ] Went to the emergency department or hospital
- [ ] Called a crisis line/mobile crisis team
- [ ] Contacted your child's psychiatrist or therapist
- [ ] Gave rescue medication
- [ ] Managed at home without outside help
- [ ] None of these

**12. Have you received training in crisis management?**

- ( ) No training
- ( ) Some guidance but no structured training
- ( ) Informal training (from therapist, videos, books, online courses)
- ( ) Formal training (e.g., Parent-Child Interaction Therapy [PCIT], Parent Management Training [PMT], Triple P, Therapeutic Crisis Intervention for Families [TCIF], Crisis Prevention Institute [CPI], Safety-Care, Handle with Care)

*[If informal or formal training selected]*

**12a. How helpful was this training for managing your child's specific behaviors?**

- ( ) 1 - Not helpful at all
- ( ) 2 - Slightly helpful
- ( ) 3 - Moderately helpful
- ( ) 4 - Very helpful
- ( ) 5 - Extremely helpful

**13. How helpful would each of these be during a crisis?** *Rate each from 1 (Not helpful) to 5 (Extremely helpful):*

Personalized strategies for my child (e.g., a plan based on my child's triggers and calming tools):

- ( ) 1 - Not helpful
- ( ) 2 - Slightly helpful
- ( ) 3 - Moderately helpful
- ( ) 4 - Very helpful
- ( ) 5 - Extremely helpful

Another person to help in the moment (e.g., trained support person or crisis coach available live):

- ( ) 1 - Not helpful
- ( ) 2 - Slightly helpful
- ( ) 3 - Moderately helpful
- ( ) 4 - Very helpful
- ( ) 5 - Extremely helpful

Emotional support for myself (e.g., check-ins, coping tips for caregivers):

- ( ) 1 - Not helpful
- ( ) 2 - Slightly helpful
- ( ) 3 - Moderately helpful
- ( ) 4 - Very helpful
- ( ) 5 - Extremely helpful

Clear step-by-step guidance (e.g., exact phrases, visuals, or prompts):

- ( ) 1 - Not helpful
- ( ) 2 - Slightly helpful
- ( ) 3 - Moderately helpful
- ( ) 4 - Very helpful
- ( ) 5 - Extremely helpful

Safety instructions (e.g., what to do first, how to keep everyone safe):

- ( ) 1 - Not helpful
- ( ) 2 - Slightly helpful
- ( ) 3 - Moderately helpful
- ( ) 4 - Very helpful
- ( ) 5 - Extremely helpful

**Progress: Section 3 Complete**
[Progress bar: 60%
[redacted]]

---

**SECTION 4: Treatment History & Access**

**14. Has your child EVER received behavioral therapy?** *Behavioral therapy means working with a therapist, counselor, or specialist to learn strategies to manage behaviors. This does NOT include medication for behaviors.*

- ( ) Yes, currently receiving
- ( ) Yes, received in the past
- ( ) Currently on waitlist
- ( ) No, due to other barriers to access
- ( ) No, never sought services

*[If "Yes, currently receiving" or "Yes, received in the past", show:]*

**14a. Which types of behavioral therapy? (check all that apply)**

- [ ] General behavioral therapy (not sure of specific type)
- [ ] Therapy for autism-related behaviors (like ABA)
- [ ] Talk therapy (like CBT or counseling)
- [ ] Family therapy
- [ ] Parent training programs
- [ ] Other: ___

**15. What barriers have you faced in accessing behavioral therapy?** (check all that apply)

- [ ] Long waitlists
- [ ] Insurance doesn't cover or limited coverage
- [ ] Therapy centers too far away
- [ ] No transportation
- [ ] Therapy times conflict with work/school
- [ ] Can't afford copays/out-of-pocket costs
- [ ] No qualified providers in my area
- [ ] Difficulty getting time off work
- [ ] No childcare for other children
- [ ] Child refused or couldn't tolerate therapy setting
- [ ] Therapist not a good fit for my child
- [ ] Language barriers
- [ ] No barriers - we've had good access
- [ ] Other: ___

**16. How many medications does your child currently take for behavioral or mental health conditions?**

- ( ) None
- ( ) 1 medication
- ( ) 2 medications
- ( ) 3 medications
- ( ) 4 or more medications

**17. Your highest education level:**

- ( ) High school or less
- ( ) Some college
- ( ) Bachelor's degree
- ( ) Graduate degree

**Progress: Section 4 Complete**
[Progress bar: 80% ████████████████████]

---

**SECTION 5: Technology Solution Preferences**

**The next questions ask what you would want from new technology.**

**18. Internet access at home: (check all that apply)**

- [ ] WiFi/Broadband
- [ ] Cellular data
- [ ] Limited/unreliable
- [ ] None

**19. What comes to mind when you hear 'artificial intelligence' or 'AI'?**

- ( ) Very familiar - I understand what it does
- ( ) Somewhat familiar - I've heard of it
- ( ) Not very familiar - I know the term but not much else
- ( ) Not familiar at all

**20. If technology could help during behavioral crises, which three features would be most valuable? Select up to three.**

[ ] Personalized crisis plan for my child

[ ] Step-by-step guidance during a crisis

[ ] Live support from a trained helper

[ ] Tools to track triggers and early warning signs

[ ] Safe de-escalation scripts and visuals

[ ] Share updates with my child’s care team

[ ] Post-crisis debrief and planning

**21. How interested would you be in the following types of technology to help you manage a behavioral crisis?**

**Phone app** *(provides guidance through your smartphone)*

- ( ) Not interested
- ( ) Somewhat interested
- ( ) Very interested

**Home system** *(cameras/speakers provide guidance without using phone)*

- ( ) Not interested
- ( ) Somewhat interested
- ( ) Very interested

**22. What concerns you MOST about technology-based crisis support?** (choose 1)

- ( ) Lack of human connection
- ( ) The technology won't understand my child's unique needs
- ( ) Privacy or data security
- ( ) Too complicated or hard to use
- ( ) Inaccurate or unhelpful recommendations
- ( ) No concerns

**23. Would you share video or audio of your child with a private, secure system if it would give you access to crisis support made just for your child? *(This is a question about future technology. We are NOT collecting video/audio in this study.)***

- ( ) Yes
- ( ) Maybe
- ( ) No

**24. Would you let your child wear a device like a smartwatch or patch that tracks things like heart rate or skin temperature to help notice early warning signs of a crisis?**

- ( ) Yes
- ( ) Maybe
- ( ) No

**Progress: Survey Complete!**
[Progress bar: 100% ████████████████████]

---

**FINAL SCREEN**

#### ✓ Thank you for completing the survey!

**Your responses will help us develop better crisis support tools for families.**

**Please click "Submit Survey" to enter the raffle for a chance to win a $25 Amazon gift card!**

**Your contact information will be stored separately from survey responses to protect privacy and will only be used for the raffle drawing.**

**Questions? Contact:**

**Raffle Entry**

### Behavioral Support Survey - Raffle Entry

Thank you for completing our survey! Enter your information below for a chance to win one of ten $25 Amazon gift cards. Your contact information will be stored separately from your survey responses to protect your privacy.

1.Name

Enter your answer

2.Email

Enter your answer

3.Phone

Enter your answer

4.Would you like to be contacted about future research studies?

Yes

No

**[SUBMIT BUTTON]**

**Thank you for completing this survey! If you entered the raffle, winners will be notified within one week after data collection closes.**

---
